# OpenMAP-T1: A Rapid Deep Learning Approach to Parcellate 280 Anatomical Regions to Cover the Whole Brain

**DOI:** 10.1101/2024.01.18.24301494

**Authors:** Kei Nishimaki, Kengo Onda, Kumpei Ikuta, Yuto Uchida, Susumu Mori, Hitoshi Iyatomi, Kenichi Oishi, the Alzheimer’s Disease Neuroimaging Initiative, the Australian Imaging Biomarkers and Lifestyle flagship study of aging

**Affiliations:** The Russell H. Morgan Department of Radiology and Radiological Science, The Johns Hopkins University School of Medicine, Baltimore, MD, USA; Department of Applied Informatics, Graduate School of Science and Engineering, Hosei University, Tokyo, Japan; The Richman Family Precision Medicine Center of Excellence in Alzheimer’s Disease, Johns Hopkins University School of Medicine, Baltimore, MD, USA

**Keywords:** Brain, MRI, T1, parcellation, segmentation, deep-learning

## Abstract

This study introduces OpenMAP-T1, a deep-learning-based method for rapid and accurate whole-brain parcellation in T1-weighted brain MRI, which aims to overcome the limitations of conventional normalization-to-atlas-based approaches and multi-atlas label-fusion (MALF) techniques. Brain image parcellation is a fundamental process in neuroscientific and clinical research, enabling a detailed analysis of specific cerebral regions. Normalization-to-atlas-based methods have been employed for this task, but they face limitations due to variations in brain morphology, especially in pathological conditions. The MALF teqhniques improved the accuracy of the image parcellation and robustness to variations in brain morphology, but at the cost of high computational demand that requires a lengthy processing time. OpenMAP-T1 integrates several convolutional neural network models across six phases: preprocessing; cropping; skull-stripping; parcellation; hemisphere segmentation; and final merging. This process involves standardizing MRI images, isolating the brain tissue, and parcellating it into 280 anatomical structures that cover the whole brain, including detailed gray and white matter structures, while simplifying the parcellation processes and incorporating robust training to handle various scan types and conditions. The OpenMAP-T1 was tested on eight available open resources, including real-world clinical images, demonstrating robustness across different datasets with variations in scanner types, magnetic field strengths, and image processing techniques, such as defacing. Compared to existing methods, OpenMAP-T1 significantly reduced the processing time per image from several hours to less than 90 seconds without compromising accuracy. It was particularly effective in handling images with intensity inhomogeneity and varying head positions, conditions commonly seen in clinical settings. The adaptability of OpenMAP-T1 to a wide range of MRI datasets and its robustness to various scan conditions highlight its potential as a versatile tool in neuroimaging.

## 1. Introduction

Brain image parcellation constitutes a pivotal aspect of neuroscientific and clinical investigations, delineating a repertoire of parcels that correspond to biologically or functionally pertinent cerebral units. These defined parcels facilitate quantitative analyses of neuroimaging data for each individual region [1–19]. While numerous criteria exist for the parcellation of cerebral territories, the designation of regional labels predominantly relies on established anatomical or neurofunctional insights. Examples include macroanatomical landmarks, such as gyri and sulci [10–12, 16, 19, 20], cellular configurations at the microscopic scale [20], the spatial distribution of transporters or receptors [21, 22], and regions characterized by vascular territories [23], as well as by functional or anatomical connectivity [5, 24–29].

The electronic version of brain atlases frequently serves as a reference for demarcating anatomical or functional territories. Such atlases typically comprise a standard brain image paired with an accompanying parcellation map, annotated with labels spanning the entirety of the cortex, white matter areas, or entire brain regions. Within atlas-based analyses, the inherent semantic information encapsulated within the parcellation map is transposed onto the target brain image for subsequent image quantification. Diverse atlas types have been formulated and employed for brain image parcellation (see [10, 16, 18, 19] for details). Image transformation techniques enable the adjustment of these knowledge-informed parcels from the atlas to conform to the specificities of the target brain. Herein, the atlas undergoes mathematical transformation (“warping” or “deformation”) to be congruent with the morphological attributes of the target brain, thus generating a parcellation map in harmony with the target’s morphology. This technique boasts over two decades of application. However, due to the pronounced individual variations in brain morphology, substantial discrepancies can sometimes be observed between the target and atlas brain structures. These mismatches make precise atlas-to-target transformations challenging [9, 30]. Notably, these transformation errors are exacerbated when addressing neuroimages of brains with pathological atrophy, lesions exhibiting signal alterations, such as ischemic or hemorrhagic lesions, or mass lesions [31]. Consequently, it has become clear that relying solely on a single brain atlas for accurate parcellation is not viable. To adeptly segment an array of brains, and encompass those that are pathologically affected, the multi-atlas label-fusion (MALF) techniques [32–39] have garnered significant traction since the 2010s.

In the MALF approach, typically 10-30 atlases are curated and subsequently transformed to the target brain. These atlases are carefully chosen to encompass a diverse range of morphological features, ensuring accommodation for inter-individual variations in cerebral morphology. This leads to the generation of as many parcellation maps as the number of atlases employed as intermediate products, each reflecting subtle differences attributable to the unique characteristics of its corresponding atlas. Leveraging these multiple parcellation outputs, a series of mathematical techniques, termed ‘label fusion,’ are employed to integrate and obtain a final optimal parcellation map for the target brain [33]. This resultant map showcases superior accuracy compared to one obtained from a single atlas [34]. Consequently, the MALF approach has found significant utility, especially in the precise parcellation of brains affected by neurodegenerative disorders that cause atrophy [39].

Given the proficiency of the MALF approach in accurately segmenting a diverse range of pathologically affected brains, its application in quantifying clinical imaging datasets that comprise various diseases seems an intuitive progression. For instance, parcellating clinical brain MRI data can yield volumetric insights into distinct cerebral regions, facilitating diagnosis, analogous image retrieval, and autonomous detection of characteristics that deviate from normative brain parameters. Nonetheless, the MALF techniques are characteristically computationally intensive, necessitating intricate mathematical transformations across multiple atlases, followed by label fusion. Consider MRICloud [38], a freely accessible cloud-based computational tool renowned for its precise brain MRI parcellation with multi-atlas label fusion. Despite the use of a cluster computing infrastructure environment, optimized for parallel processing, the use of the computationally intensive advanced large deformation diffeomorphic metric mapping for image transformation results in several hours of processing time for a single image. This computational burden poses significant challenges in big-data analysis or clinical scenarios. For instance, in clinical settings where multiple brain MRI scans necessitate immediate segmentation for diagnostic assistance or when a large repository of brain images requires quantification, expedited processing is imperative. Under such circumstances, the current MALF techniques prove overly resource-intensive and time-prohibitive.

In recent years, there has been a growing inclination to use deep-learning models to expedite the parcellation of brain MRI while simultaneously enhancing accuracy [40–47]. For instance, ParcelCortex [45] employs convolutional neural networks (CNNs), while DeepParcellation [46] integrates the Attention 3D U-Net for cortical segmentation based on the Desikan-Killiany-Tourville atlas parcellation derived from FreeSurfer [48]. A salient advantage of deep-learning lies in its efficiency: once a model is trained and validated, it permits rapid parcellation. This proficiency renders it advantageous for processing extensive datasets, encompassing both research-oriented and clinical imaging. Nonetheless, models capable of detailed segmentation across all cerebral regions, inclusive of white matter territories, and those offering results comparable to the sophisticated MALF method, remain underdeveloped.

In the present study, we introduce a deep-learning-based, rapid, whole-brain parcellation method. Our aim was to construct a model that not only mirrors the accuracy of the multi-atlas approach, but also accommodates images from diverse repositories and facilitates parcellation within mere minutes on a standard desktop configuration. The model has been named “Open resource for Multiple Anatomical structure Parcellation for T1-weighted brain MRI (OpenMAP-T1)” and is accessible through the website (URL: https://github.com/OishiLab/OpenMAP-T1).

## 2. Materials and Methods

### 2.1. Participants

We used public brain MRI datasets for the model training and evaluations: Alzheimer’s Disease Neuroimaging Initiative 2 and 3 (ADNI2/ADNI3) [49], Australian Imaging, Biomarkers and Lifestyle (AIBL) [50, 51], Calgary-Campinas-359 (CC-359) [52], LONI Probabilistic Brain Atlas (LPBA40) [53], Neurofeedback Skull-stripped (NFBS) [54], and Open Access Series of Imaging Studies 1 and 4 (OASIS1/OASIS4) [55, 56]. Table 1 shows the dataset descriptions used in this study. Three-hundred-fifty baseline MRIs of ADNI2 were used to train the OpenMAP-T1, and other MRIs, including 750 ADNI2. MRIs not used to train the model were used to test the model. Although these datasets include multiple scans from single participants, only one MRI per participant was randomly selected to avoid potential bias toward specific individuals.

**Table 1.** Dataset used in our study: Alzheimer’s Disease Neuroimaging Initiative 2/3 (ADNI2/ADNI3), Australian Imaging, Biomarkers and Lifestyle (AIBL), Calgary-Campinas-12 (CC-12), LONI Probabilistic Brain Atlas (LPBA40), Neurofeedback Skull-stripped (NFBS) Repository, Open Access Series of Imaging Studies 1/4 (OASIS1/OASIS4). OASIS4 consists of clinical MRIs with various diseases and conditions. OpenMAP-T1 was trained using only 350 cases in ADNI2. AD, Alzheimer’s disease; MCI, mild cognitive impairment; CN, cognitively normal older people. One MRI per subject was randomly selected to avoid potential bias.

|  |  | Train |  |  |  | Test |  |  |  |  |
| --- | --- | --- | --- | --- | --- | --- | --- | --- | --- | --- |
| Dataset |  | ADNI2 | ADNI2 | ADNI3 | AIBL | CC359 | LPBA40 | NFBS | OASIS1 | OASIS4 |
| # Subjects |  | 350 | 750 | 929 | 376 | 359 | 40 | 125 | 235 | 570 |
| # Voxels<br>($mm^3$) | | 1.05~1.43 | 1.05~2.18 | 0.99~1.44 | 1.19~1.20 | 0.79~1.33 | 0.91~1.10 | 0.99 | 1.25 | 0.22~1.52 |
| Age | mean | 73.7 | 74.3 | 73.7 | 74.0 | 53.4 | 29.2 | 31.0 | 72.3 | 72.5 |
|  | std | 7.4 | 7.5 | 8.0 | 7.1 | 7.8 | 6.3 | 6.6 | 12.0 | 9.2 |
| Sex | F | 160 | 353 | 486 | 198 | 183 | 20 | 77 | 156 | 303 |
|  | M | 190 | 397 | 443 | 178 | 176 | 20 | 48 | 79 | 267 |
| Manufacturer | GE | 97 | 254 | 211 |  | 120 | 40 |  |  |  |
|  | PHILIPS | 65 | 126 | 142 |  | 119 |  |  |  |  |
|  | SIEMENS | 188 | 370 | 576 | 376 | 120 |  | 125 | 235 | 570 |
| Field Strength | 1.5T | 30 | 149 |  | 102 | 179 | 40 |  | 235 | 29 |
|  | 3T | 320 | 601 | 929 | 274 | 180 |  | 125 |  | 541 |
| Diagnosis | CN | 122 | 259 | 522 | 268 | 359 | 40 | 125 | 135 |  |
|  | MCI | 196 | 318 | 301 | 64 |  |  |  |  | Real-World MRI |
|  | AD | 32 | 173 | 106 | 44 |  |  |  | 100 |  |

The ADNI [49] was launched in 2003 as a public-private partnership, led by Principal Investigator Michael W. Weiner, MD. The primary goal of ADNI has been to test whether serial magnetic resonance imaging (MRI), positron emission tomography (PET), other biological markers, and clinical and neuropsychological assessment can be combined to measure the progression of mild cognitive impairment (MCI) and early Alzheimer’s disease (AD). For up-to-date information, see www.adni-info.org. The ADNI study has evolved through several phases, with ADNI2 during 2011 - 2016 and ADNI3 during 2016 - 2022 being two of them. For our study, we have included Magnetization Prepared Rapid Acquisition Gradient Echo (MPRAGE) images from ADNI2 and ADNI3 datasets. As recommended by the ADNI team, we included ADNI2 MPRAGE images (ADNI2: 55.1-94.7 years, ADNI3: 50.5-97.4 years) preprocessed with Gradwarp, B1 non-uniformity, and N3 bias field corrections. Note that the preprocessing was not required for the ADNI3 MPRAGE images since, such corrections are internally applied by individual vendors.

The AIBL [50, 51], also known as Australian ADNI, is a long-term research initiative that aims to understand the biomarkers and cognitive characteristics that determine the development of AD. The AIBL study commenced in 2006 and the methodology has been reported previously [50]. In our study, the the original MPRAGE images (55.0-96.0 years) downloaded from the website (https://aibl.org.au/) were included.

The CC359 [52] dataset is an open, multi-vendor, multi-field-strength brain MRI dataset. It is composed of 359 MRIs of healthy adults acquired on scanners from three vendors (Siemens, Philips, and General Electric (GE)) at both 1.5 T and 3 T to evaluate the impact of scanner vendor and magnetic field strength on skull-stripping. In our study, we used MPRAGE images of Siemens and Philips and 3D spoiled gradient echo sequence (SPGR) (29.0-80.0 years) on the GE, downloaded from the website (https://www.ccdataset.com/download).

The LPBA40 [53] is a set of brain MRIs of 40 healthy young adults scanned on a single 1.5T GE scanner. In our study, we used the 3D SPGR images (19.3-39.5 years) downloaded from the website (https://www.loni.usc.edu/research/atlas_downloads).

The NFBS [54] dataset is a repository of brain MRIs of 125 individuals, including 66 who were diagnosed with a wide range of psychiatric disorders, scanned on a single 3T Siemens scanner. In our study, we used MPRAGE images (21.0-45.0) downloaded from the website (http://preprocessed-connectomes-project.org/NFB_skullstripped/).

The OASIS [55, 56] is a brain MRI dataset that includes multiple releases, such as OASIS-1, OASIS-2, OASIS-3, and OASIS-4, which provide cross-sectional and longitudinal MRI data for normal aging and AD. The subjects of the OASIS1 were selected from a larger database of individuals who had participated in MRI studies at Washington University. OASIS4 is a Clinical Cohort and was acquired at the Memory Diagnostic Center. All subjects in OASIS4 underwent a clinical assessment conducted by experienced clinicians. The 570 subjects we selected included 16 different diagnostic labels. In our study, we used MPRAGE images (OASIS1: 33.0-96.0, OASIS4: 37.0-94.0) downloaded from the website (https://www.oasis-brains.org/).

### 2.2. Anatomical Labeling for Training and Evaluation

The parcellation map obtained from the MALF algorithm [57, 58] and implemented in MRICloud (www.MRICloud.org Johns Hopkins University, Baltimore, MD, USA) [38] was used to generate anatomical labels for model training and evaluation. The MALF algorithm parcellates the entire brain into 280 brain regions. We went over the 350 MRIs used as a training dataset to confirm that there was no substantial mislabeling; therefore, manual correction was not performed. Details of the labels used to train OpenMAP-T1 are shown in Figure A in the Supplementary Material. It should be noted that, in this paper, the term ‘MALF’ will henceforth refer exclusively to the specific algorithm implemented in MRICloud.

### 2.3. Model Design

The OpenMAP-T1 was designed to accept any T1-weighted images and output a corresponding parcellation map, in which gray matter, white matter, and cerebrospinal fluid areas are segmented and further parcellated into 280 anatomical regions based on the JHU-atlas [10].

Figure 1 shows the overview of OpenMAP-T1. The OpenMAP-T1 follows six phases: (1) preprocessing; (2) application of a cropping network (CNet) consisting of the 2D U-Net [59] to crop the area surrounding the head in the input MRI; (3) application of a skull-stripping network (SSNet) consisting of the 2D U-Net to extract the brain; (4) application of a parcellation network (PNet) consisting of the 2.5D U-Net [60] to parcellate the whole brain into 141 anatomical areas; (5) application of a hemisphere network (HNet) consisting of 2D U-Net to segment the whole brain into the right and left hemisphere; and (6) separation of 139 regions of the 141 from the parcellation map created in phase (4) into right and left sides based on the hemisphere map from phase (5), with the exception of the 3rd and 4th ventricles. Consequently, OpenMAP-T1 produces a parcellation map consisting of 280 neuroanatomically defined regions.

**Figure 1.**
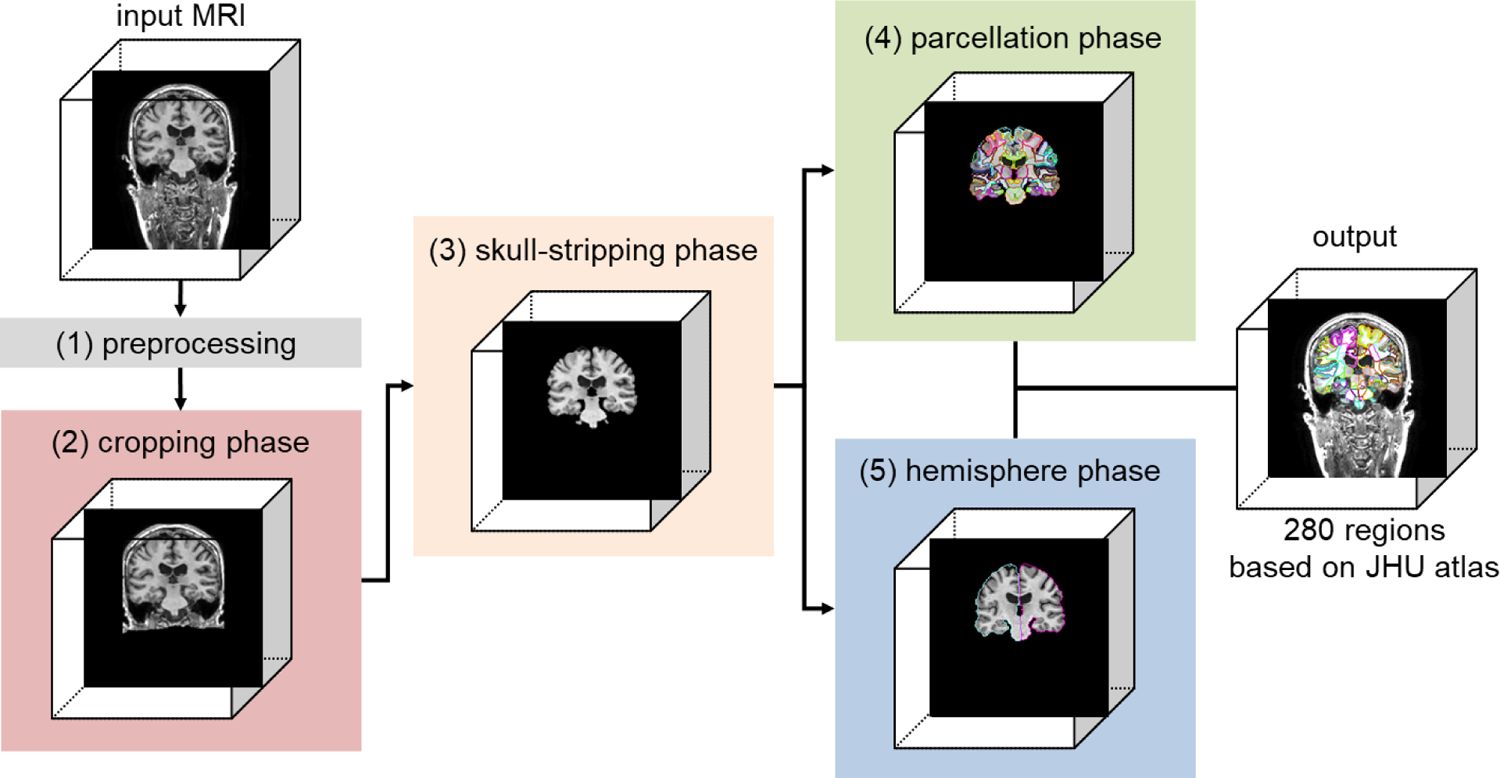
Overview of the open resource for multiple anatomical structure parcellation for T1-weighted brain MRI (OpenMAP-T1) consisting of the preprocessing, cropping phase, skull-stripping phase, parcellation phase, and hemisphere phase.

#### 2.4.1. Preprocessing

N4 bias field correction [61] was applied to remove intensity non-uniformity. In addition, the images were rescaled to a resolution of 1 × 1 × 1 mm using trilinear interpolation, and their size was standardized to 256 × 256 × 256 through the application of zero padding. Pixels with intensity values below 0 or above u+2σ (where u is the mean and σ is the standard deviation) were identified as outliers and excluded. These pixels were then linearly normalized to a range between −1 and 1. Following normalization, the excluded pixels were replaced with the minimum and maximum values within this normalized range.

#### 2.4.2 Cropping phase

Non-brain soft tissue, such as skin, fat, and muscle, potentially confound whole-brain parcellation. In particular, variations in the intensity of the neck tissue interferes with image segmentation. In addition, the extent to which the field-of-view of the image covers the neck depends on the scan parameters, and, in some publicly available datasets (e.g., NFBS and OASIS1), a defacing algorithm was applied to the image for de-identification [54, 55]. To reduce the influence of neck tissue in image segmentation and parcellation, we set the cropping phase to eliminate the regions below the brain in a consistent way using the CNet. Figure B in the Supplementary Materials shows an overview of the cropping phase and the detail of the CNet is shown in Section 2.4.6.

In the training of the CNet, the head region included in the parcellation map resulting from MALF was used (Figure A in the Supplementary Material), since the parcellation map covers the head above the foramen magnum, and does not include neck tissue below it. The CNet employed 2D segmentation on individual cross-sections of a 3D brain MRI, which were then vertically stacked for high-speed processing. By utilizing the 2D U-Net architecture, it was possible to train the model using numerous images from a single MRI. For example, with a 256×256×256 matrix, 2D U-Net can utilize 256 slices from a single MRI. Note that the cropping phase was specifically designed to eliminate tissue located below the brain; hence, the axial section, which does not contain vertical information, was excluded from the ensemble. The output is a probability map for the head above the foramen magnum, and areas with a probibability of 50% or higher are defined as head masks. A failure in the cropping phase considerably affects the processing performance of subsequent phases. To prevent small gaps or missing areas in the output mask, a closing process was applied using a 3 × 3 × 3 filter. The dilation and erosion operations were performed three times each.

#### 2.4.3. Skull-stripping phase

To further remove signals from extracranial soft tissues that remain after the initial cropping phase, we applied skull-stripping to the image. Given the varying intensity ranges caused by different scanner types, scan sequences, and parameters, it is essential to adjust the image contrast between gray and white matter, as well as cerebrospinal fluid. However, signals from extracranial soft tissues, such as fat, bone, and muscle, vary greatly and can disrupt the stable signal intensity profile of intracranial structures. This variation can adversely affect the performance of deep-learning models. By employing skull-stripping with SSNet, we effectively removed irrelevant regions for the later stages of parcellation and hemisphere analysis. Figure C in the Supplementary Materials provides an overview of the skull-stripping phase and the detail of the SSNet is shown in Section 2.4.6.

To develop SSNet, we trained the model using an intracranial mask obtained from MALF (Figure A in the Supplementary Material). SSNet performs 2D segmentation on any cross-sectional view of a 3D brain MRI, stacking these sections vertically in a manner similar to that of CNet. We input three cross-sectional views—sagittal, coronal, and axial—into a 2D-U-Net. The output is a label for the intracranial space, defined as areas with a probability of 50% or higher.

#### 2.4.4. Parcellation phase

In our current computing environment, training a full-size deep-learning network for multi-class parcellation with 3D images, such as an entire brain, presented challenges. To optimize parcellation performance without running GPU memory, we used two strategies: utilizing 2D slices, and merging the left and right brain regions. An overview of the parcellation phase can be seen in Figure D of the Supplementary Materials.

The PNet, designed for 2D segmentation of 3D brain MRI cross-sections, stacks these sections vertically. It is important to note that the PNet functions as a 2.5D U-Net, incorporating a target slice and the slices directly above and below it, and combining these slices along the channel direction. In typical 2D U-Net applications for 3D images, vertical spatial information is lost. However, the 2.5D U-Net approach allows for the use of pseudo spatial information while reducing parameter count compared to a 3D U-Net. Consequently, for each cross-section, three image slices (256 x 256) are stacked in the depth direction, resulting in an input dimension of 256 x 256 x 3 for the PNet. The choice to stack three slices was based on the findings from preliminary experiments.Furthermore, of the 280 brain regions, those present in both hemispheres were merged as a single region (Figure A in the Supplementary Material).

Excluding the 3rd and 4th ventricles, 278 regions exist in both the left and right hemispheres, as per the JHU-atlas. During the parcellation phase, this merging reduces the target regions for PNet to 141, making the segmentation task effectively 142 classes, including the background.

Combining the left and right regions offers three benefits. First, it enables U-Net to train on larger regions, which is crucial as some of the 280 regions are smaller than 100 voxels and challenging to extract accurately. Merging hemispheres roughly doubles the volume of these regions, making them more tractable. Second, it reduces computational costs. In 2D or 2.5D U-Net, the dimension of the output parcellation map is height × width × channels (number of segmentation classes). By merging hemispheres, we halve the number of output channels. Third, it facilitates the use of sagittal sections. Normally, distinguishing left from right in sagittal sections is tough, and can potentially degrade parcellation performance. This issue is resolved by treating the hemispheres as identical.

Unlike CNet and SSNet, PNet uses three models for each cross-section (coronal, sagittal, axial), and the final prediction is based on the highest average prediction probability across these models. This approach was chosen to ensure consistent predictions in regions where parcellation is challenging, such as at the brain’s edges. Training a single model with three cross-sections risks misidentifications, like mistaking a coronal for an axial section.

#### 2.4.5. Hemisphere phase

The segmentation task of HNet is a three-class classification: background; right hemisphere; and left hemisphere. This classification aims to determine the left and right borders of 139 of the 141 region labels generated in the parcellation phase. The hemisphere phase model was trained using hemisphere labels obtained from MALF. An overview of this phase is depicted in Figure E of the Supplementary Materials and the detail of the HNet is shown in Section 2.4.6.

HNet employs 2D segmentation on any cross-section, stacking these sections vertically. It specifically uses axial and coronal sections to output the hemisphere labels. A post-processing step involving dilation was implemented to separate each region generated during the parcellation phase to the right and left sides. The process is as follows: (1) Dilate only the left hemisphere label. (2) Modify the overlapping parts of the dilated left hemisphere label with the right hemisphere label to be classified as right side. (3) Dilate only the right hemisphere label. (4) Modify the overlapping parts of the dilated right hemisphere label with the left hemisphere label to be classified as left side. This method allows the hemisphere labels to expand while minimally impacting the borders.

Finally, the parcellation map, which includes 280 region labels, was created by dividing the 141-region parcellation map into left and right hemispheres, using the hemisphere labels obtained from the hemisphere phase. It is important to note that in the 141-region parcellation map, the two regions without a left-right distinction (3rd and 4th ventricles) were given priority over the hemisphere labels (i.e., the hemisphere label was ignored). Furthermore, if there was an overlap between the background from the parcellation phase and the hemisphere region from the hemisphere phase, the background was given precedence.

#### 2.4.6. Structures of CNet, SSet, PNet, and HNet and their training

The CNet, SSNet, PNet, and HNet each comprise 24 convolution layers based on 2D CNN (Figure 2). The main distinction between them lies in the number of input and output channels. As detailed in Figure 2, CNet, SSNet, and HNet had one input channel, while PNet had three channels.

**Figure 2.**
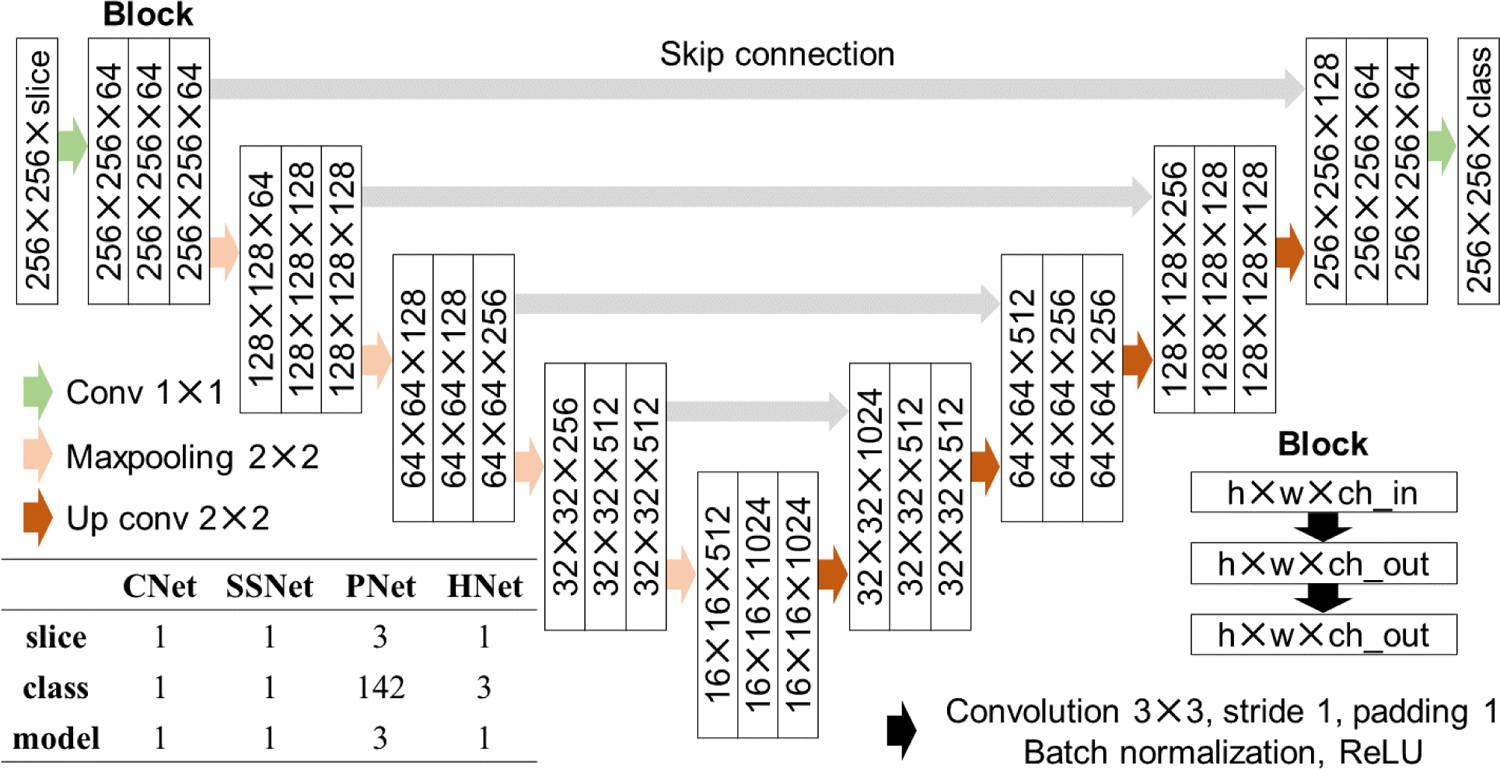
Architecture of the U-Net (CNet, SSNet, PNet, and HNet). U-Net consists of nine blocks, and each block includes two convolutions, batch normalization, and a ReLU function. The CNet, SSNet, and HNet have one input channel, and only PNet has three channels, including the target slice and its upper and lower slices. Since the CNet and SSNet perform binary cross entropy, the output class is only one. Also, since the PNet and HNet use multi-class classification, the output is the number of regions plus one (background). Note that only PNet has three models (for coronal, sagittal, and axial sections).

Data augmentation included a random rotation ranging from −20 to +20 degrees and a random shift from −20 to +20 pixels, applied with an 80% probability. For the sagittal cross-sections, a broader random rotation range of −30 to +30 degrees was used, based on the greater variation seen on existing MRI databases. To ensure robust performance for low signal-to-noise ratio (SNR) images, random Gaussian noise (mean 0, standard deviation 0.25) was added with a 50% probability. These data augmentation techniques were applied consistently across all phases.

MRI magnetic field inhomogeneity led to very low-frequency intensity variations throughout the image. Consequently, a random bias field [62] was introduced with a 50% probability during the cropping and skull-stripping phases. While N4 bias field correction was used in preprocessing to address inhomogeneity as much as possible, this data augmentation was implemented to enhance the model’s robustness. To adapt the CNet for use with images that did not include the neck area below the foramen magna, commonly found in axial scans and defaced images, we augmented the training data by incorporating images with the neck area cropped out, at a probability of 20%. Similar to CNet, the training dataset for SSNet was augmented by including skull-stripped images as inputs with a 20% probability. This approach enables the model to more accurately identify the location of the brain by reducing the impact of non-brain regions.

The CNet, SSNet, PNet, and HNet were mainly trained using two RTX 3090 graphics processing units (GPU) with 24GB memory for about 24 hours. Also, Automatic Mixed Precision (AMP) was utilized to accelerate training. The number of epochs was set to 10,000, and the learning rate was set to decrease sequentially from 0.01 to 0.0001 by the Cosine Annealing Learning Rate Scheduler. The batch size was 64 on CNet, SSNet, and HNet, and 32 on PNet. For the loss functions, the CNet and SSNet used binary cross-entropy, while the PNet used a combination of cross-entropy and Dice loss. The HNet exclusively used cross-entropy.

### 2.5. Evaluation of parcellation performance

We assessed the effectiveness of OpenMAP-T1 in terms of technical and biological criteria. The technical assessment encompassed three areas: parcellation performance; generalizability; and processing time. For the biological evaluation, OpenMAP-T1 was tested under the assumption of its application in AD research. Furthermore, we compared the parcellation accuracy of OpenMAP-T1 with that of FreeSurfer (version 7.4.1), a widely used tool in brain MRI parcellation.

#### 2.5.1. Technical evaluation

##### Parcellation performance

To evaluate the parcellation performance of OpenMAP-T1, four metrics were utilized: (1) illustrate the spatial overlap between two parcellation maps generated by MALF and OpenMAP-T1 by calculating the average recall and precision, considering the results of MALF as the ground truth, and also compute the Dice score; (2) establish correlation analyses of the predicted volumes between MALF and OpenMAP-T1, indicating the consistency of the relationship between these predicted volumes, with both Pearson and Spearman correlation coefficients calculated; (3) use a Bland-Altman plot [63] to investigate agreement between regional volumes predicted by MALF and OpenMAP-T1, allowing identification of systematic bias between the measurements and outliers; and (4) analyze the relationship between the volume predicted by MALF and the corresponding Dice score, showing how the predicted volume correlates with parcellation performance. These metrics were determined for each individual region.

##### Generalizability

For parcellation methods to be effectively applicable in large-scale clinical research, they must possess a high degree of generalizability. To assess the generalizability of OpenMAP-T1, we examined whether significant variations in the Dice score were present based on factors such as database, scanner manufacturer, magnetic field strength, age, sex, and diagnosis. We employed analysis of variance (ANOVA) for this comparative analysis. A p-value less than 0.05 in two-sided tests was deemed to reflect statistical significance.

#### 2.5.2. Comparison with FreeSurfer

It is important to acknowledge that a direct comparison between the parcellation maps of OpenMAP-T1, which uses the JHU-atlas [10], and FreeSurfer, based on the Desikan-Killiany-Tourville (DKT) atlas [18], is not feasible because their anatomical definitions are different.

Consequently, we conducted a focused comparison of OpenMAP-T1 and FreeSurfer, specifically examining the hippocampus, amygdala, and entorhinal cortex, as these regions are notably associated with Alzheimer’s Disease (AD). Two metrics were used for this comparison: (1) the average recall, precision, and Dice score for the parcellation maps of the hippocampus, amygdala, and entorhinal cortex produced by OpenMAP-T1 and FreeSurfer, considering the result of FreeSurfer as the ground truth; (2) a correlation analysis of the predicted volumes for the hippocampus, amygdala, and entorhinal cortex, comparing the results from OpenMAP-T1 and FreeSurfer.

#### 2.5.3. Biological evaluation

We assessed the diagnostic ability to distinguish between AD and CN participants using predicted volumes derived from MALF, OpenMAP-T1, and FreeSurfer. This evaluation involved 628 subjects with AD or CN labels from the ADNI3 dataset, using a three-fold cross-validation approach. A logistic regression model with Least Absolute Shrinkage and Selection Operator (LASSO) was utilized for the classification. To mitigate multicollinearity, the predicted volumes of 280 anatomical structures by MALF and OpenMAP-T1 were averaged across their left and right counterparts. Similarly, the volumes calculated by FreeSurfer for bilateral regions were also averaged. In addition, we adjusted for brain size effects by normalizing each structure’s volume to the total brain volume of each participant. These normalized volumes were then transformed into z-scores, based on the mean and standard deviation, and used as input variables for the model. The classification performance was compared using the Area Under the Curve (AUC) from receiver operating characteristic (ROC) curve analysis, with significant differences determined using DeLong’s algorithm [64]. A p-value less than 0.05 in two-sided tests was considered statistically significant. Furthermore, we identified the top 20 structures based on the coefficients of the LASSO model after training to understand the important regions of this model.

## 3. Results

### 3.1. Technical evaluation

#### Parcellation Performance

Figure 3 presents a comparative analysis of parcellation results between MALF and OpenMAP-T1. Despite significant variations in head appearance across different datasets, OpenMAP-T1 demonstrated satisfactory performance for all eight datasets. Notably, the median Dice score exceeded 0.8 for every dataset, with the exception of LPBA40, which did not utilize MPRAGE imaging.

**Figure 3.**
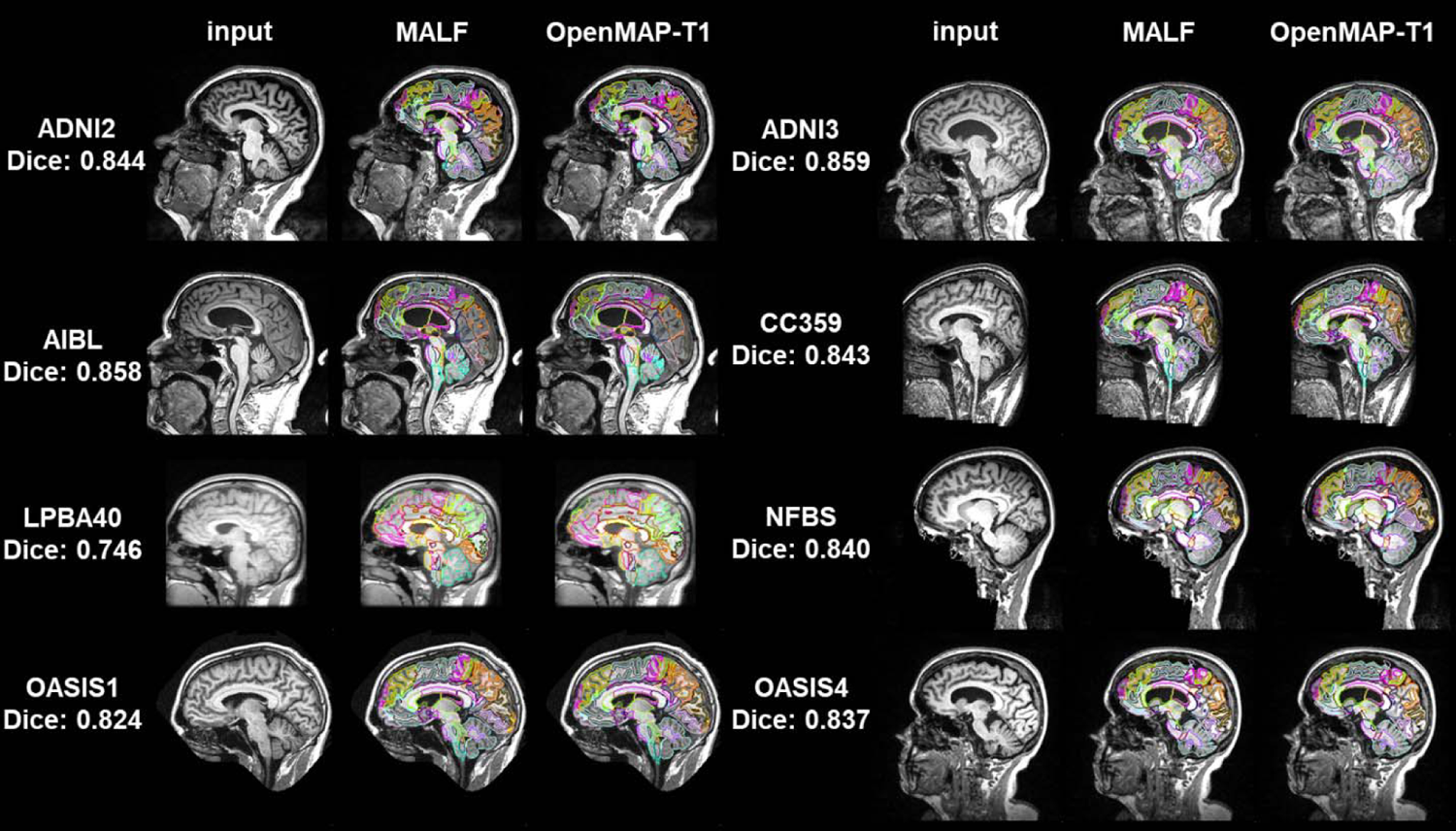
Representative results from MALF and OpenMAP-T1 are demonstrated. The median Dice score calculated between the two sets of results is provided. Note that defacing techniques have been implemented on NFBS and OASIS1 datasets to safeguard the privacy of the participants.

Figure 4 illustrates the average recall, precision, and Dice scores obtained from comparing 280 anatomical regions in the parcellation maps between MALF and OpenMAP-T1, considering the results of MALF as the ground truth. These scores, exceeding 0.75 in all datasets, signify a considerable level of agreement between the two methods. However, it should be noted that some datasets displayed extreme outliers.

**Figure 4.**
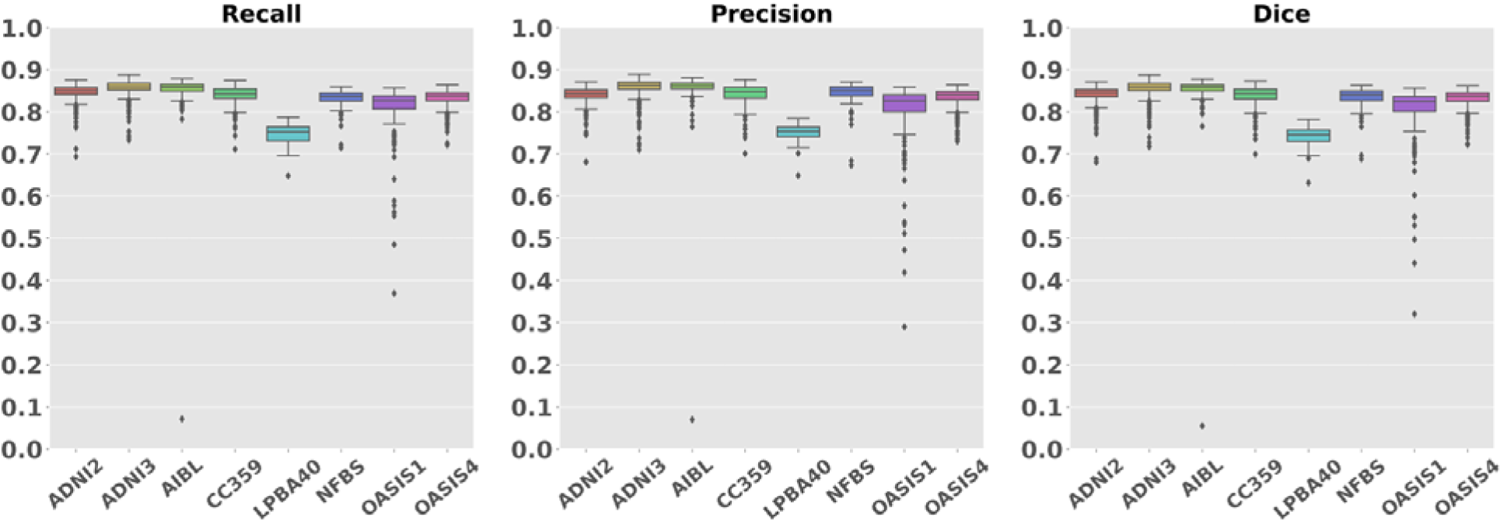
Boxplot of the recall, precision, and Dice scores across each dataset.

Figure 5 shows the image with the lowest Dice score and its underlying causes. The causes were predominantly attributed to mislabeling by MALF, except for one image from OASIS4. In the image from ADNI2, ROIs for gray and white matter in the cerebellum erroneously included neck areas. In the image from ADNI3, a portion of the occipital lobe was absent from the parcellation map. The image from AIBL was affected by extremely high-intensity noise, leading to significant mislabeling in the MALF results. In the image from CC359, the labels extended beyond the parietal lobe boundaries. In images from LPBA40 and OASIS1, parts of the cerebellum were missing in the parcellation map, a pattern also observed in other OASIS1 images. The image from NFBS had an issue with a missing section of the frontal lobe in its parcellation map. The exception was an image from OASIS4 with a large arachnoidal cyst that neither method successfully labeled.

**Figure 5.**
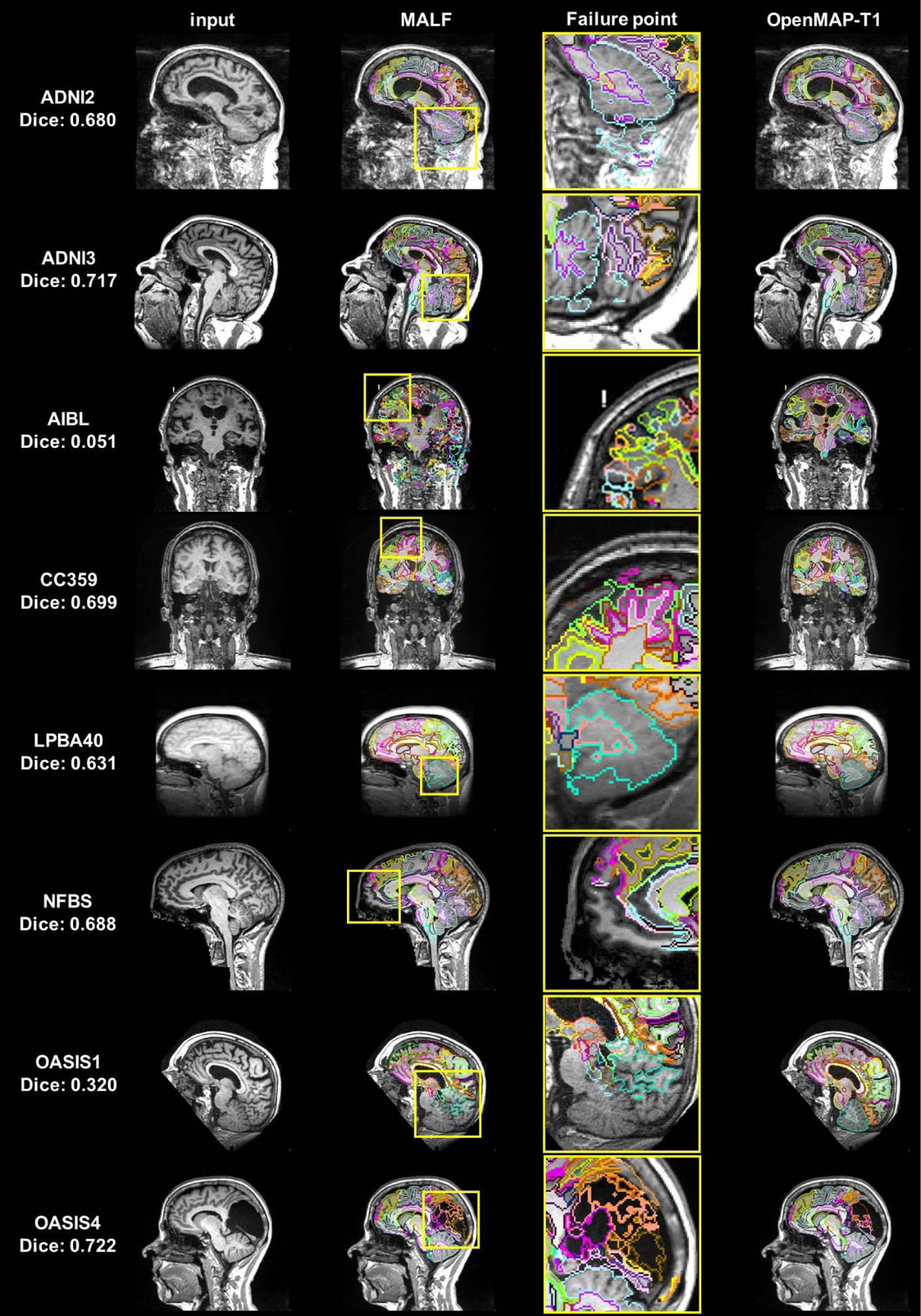
Image with the lowest Dice scores from all datasets (From left to right: input image, output of MALF, enlarged image of MALF failure point, output of OpenMAP-T1). The yellow squares on the images in the second column correspond to the enlarged images in the third row and highlight areas where the MALF’s parcellation failed.

Figure 6A presents a comparison of the region volumes predicted by MALF and OpenMAP-T1 for the ADNI3 and OASIS4 databases. The correlation coefficients for these comparisons were above 0.99 using both Pearson and Spearman methods, indicating an almost perfect correlation. The Bland-Altman plot revealed that the majority of regions had errors within the 2σ range, demonstrating excellent agreement between MALF and OpenMAP-T1. However, a few regions in both the ADNI3 and OASIS4 databases exhibited disagreements exceeding 2σ. Figure 6C shows a weak correlation between the volumes of the regions and their Dice scores (R = 0.660 in ADNI3, R = 0.707 in OASIS4), despite most regions having a Dice score higher than 0.7. This implies that smaller regions tend to have greater discrepancies between the measurements from MALF and OpenMAP-T1. Additional results for other databases can be found in the Supplementary Material (Figure F, G and H).

**Figure 6.**
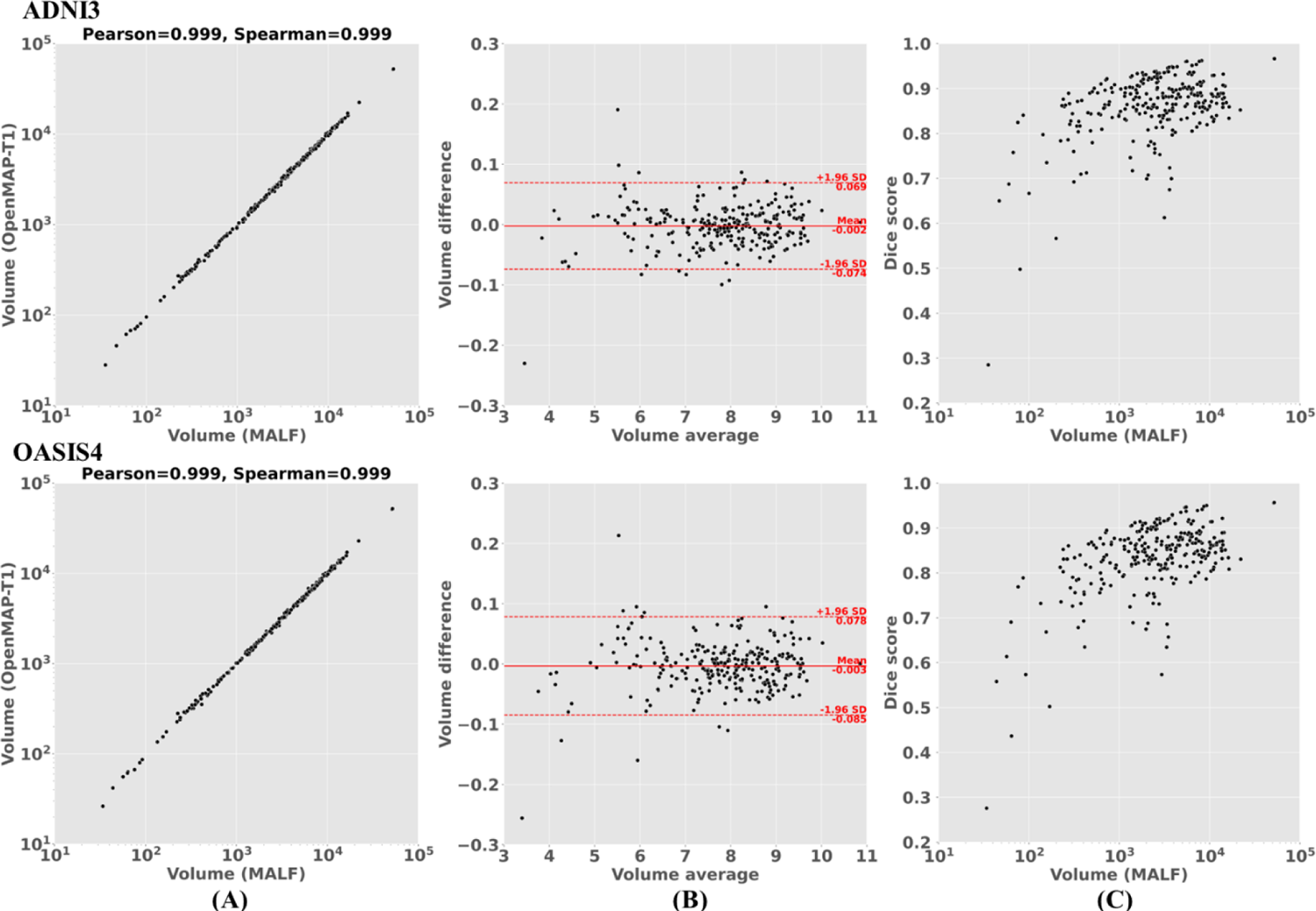
(A) Correlation between the predicted volumes obtained using MALF and OpenMAP-T1. (B) Bland-Altman plot to demonstrate agreement between regional volumes predicted by MALF and OpenMAP-T1. The volume measurements were transformed using a base-2 logarithmic scale. (C) Relationship between the structural volume obtained from MALF and the Dice score between MALF and OpenMAP-T1.

We noted that certain anatomical labels were absent from the predicted parcellation map produced by OpenMAP-T1, specifically in the right rostral anterior cingulate white matter and the bilateral fimbria. A similar trend of omitting these small structures was observed with MALF. MALF results were as follows: 1.26 mm³ with 924 of 929 instances for the right rostral anterior cingulate white matter; 6.37 mm³ with 493 of 929 instances for the left fimbria; and 12.86 mm³ with 136 of 929 instances for the right fimbria.

#### Generalizability

Figure 7 shows the Dice scores across biological (age, sex, and diagnosis) and technological (scanner manufacturer and field strength) categories, for the ADNI3 and OASIS4 datasets. The results from other databases are provided in the Supplementary Material (Table A). The red boxplots illustrate the influence of biological factors, while the blue boxplots represent the impact of technological factors. The Dice scores exceeded 0.8 in all conditions. Age had a significant effect in ADNI2, where the Dice score tended to decrease in individuals in their 90s. A similar trend was observed in OASIS4, although the effect was not statistically significant. No significant differences were noted based on sex in either database. Regarding the diagnosis, a significant effect was observed in OASIS4, where a diagnosis of VCI was associated with lower Dice scores compared to other diagnoses; however, in ADNI3, the diagnosis did not significantly impact Dice scores. Among the scanner manufacturers, GE systems showed lower Dice scores compared to those from Philips and Siemens. In terms of field strength, scanners operating at 1.5 T had lower Dice scores compared to those operating at 3T.

**Figue 7.**
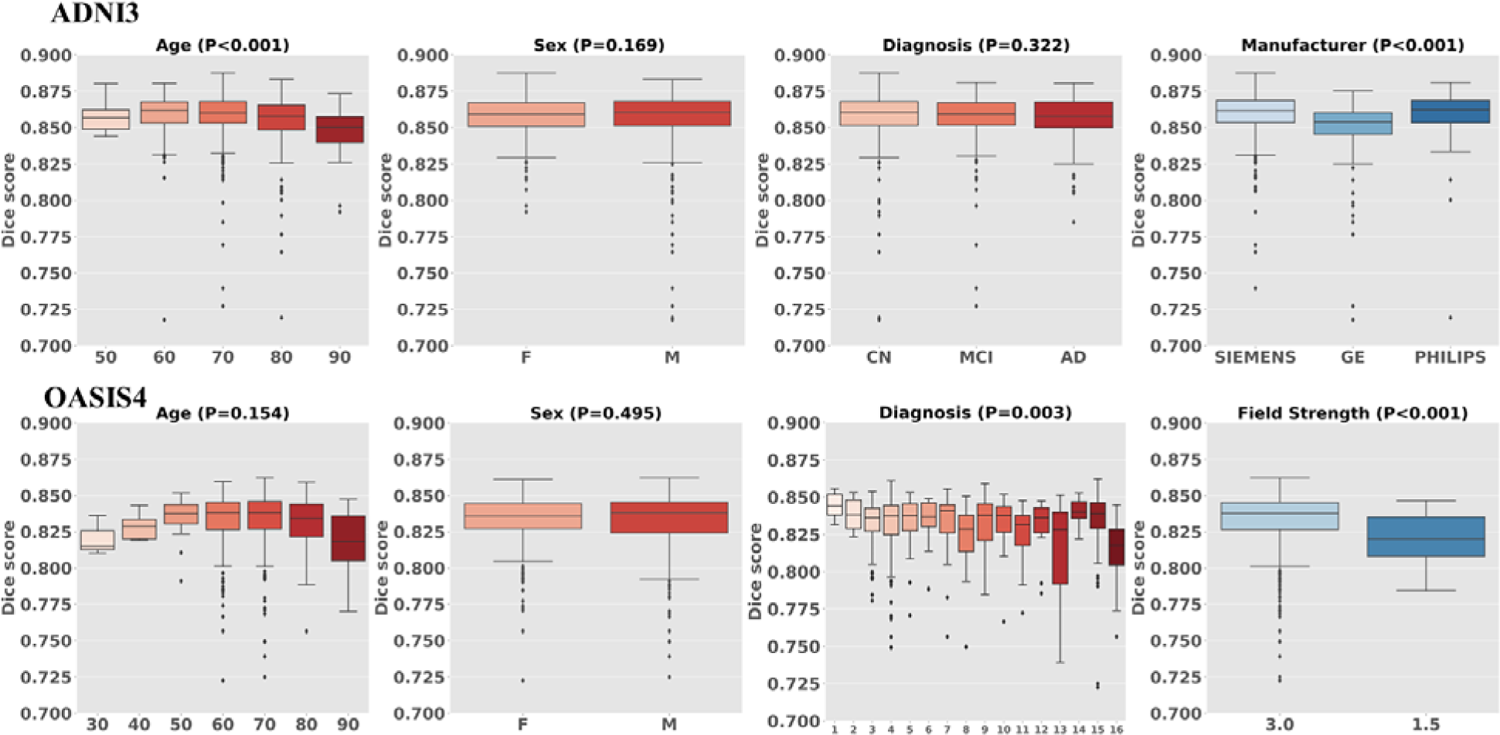
Boxplot of biological (age, sex, diagnosis) and technological effects (manufacturer, field strength) with Dice score. OASIS4 includes 16 diagnostic labels: 1: AD variant; 2: AD + non neurodegenerative; 3: AD / vascular; 4: alzheimer disease dementia; 5: Cognitively Normal (CN); 6: Dementia with Lewy Bodies (DLB); 7: early onset AD; 8: Frontotemporal Dementia (FTD); 9: Mild Cognitive Impairment (MCI); 10: mood / polypharmacy / sleep; 11: non-neurodegenerative neurologic disease; 12: other-miscellaneous; 13: other non-AD neurodegenerative disorder; 14: Primary Progressive Aphasia (PPA); 15: uncertain - AD possible; 16: Vascular Cognitive Impairment (VCI).

#### Runtime

OpenMAP-T1 performed complete parcellation at 90 sec/case using a single GPU (RTX3090) and 10 min/case using a CPU (i9-10980XE). This result shows that OpenMAP-T1 is 40 times faster than MRICloud, which is one hour/case.

### 3.2. Comparison with FreeSurfer

Figure 8 shows the recall, precision, and Dice scores for the hippocampus, amygdala, and entorhinal cortex in the parcellation maps of OpenMAP-T1 and FreeSurfer, considering the results of FreeSurfer as the ground truth. Figure 9 compares the predicted volumes in the hippocampus, amygdala, and entorhinal cortex between OpenMAP-T1 and FreeSurfer. In the hippocampus, the recall, precision, and Dice scores were all high. This result indicated that the definition of the hippocampus boundary was nearly identical in OpenMAP-T1 and FreeSurfer, and both methods were equally accurate in identifying the hippocampus. As shown in Figure 9, the hippocampal volume measurements obtained by both methods correlated well. In the amygdala, although the recall was equivalent to that of the hippocampus, the precision and Dice scores were lower. This suggests the existence of systematic bias due to methodological differences in defining the amygdala region; the amygdala region defined by FreeSurfer was smaller than that defined by OpenMAP-T1, which includes the cortical amygdala not included in the FreeSurfer definition. As demonstrated in Figure 9, although the volume measurements of both methods correlated well, FreeSurfer consistently underestimated the amygdala volume compared to OpenMAP-T1. Conversely, FreeSurfer defined the entorhinal cortex area as larger than OpenMAP-T1 and often includeed the adjacent dura matter within the entorhinal cortex region (see Figure 10). Therefore, while precision was relatively preserved, the recall and Dice scores were lower, and the correlation of volume measurements between the two methods was weaker for the entorhinal cortex compared to the hippocampus and amygdala.

**Figure 8.**
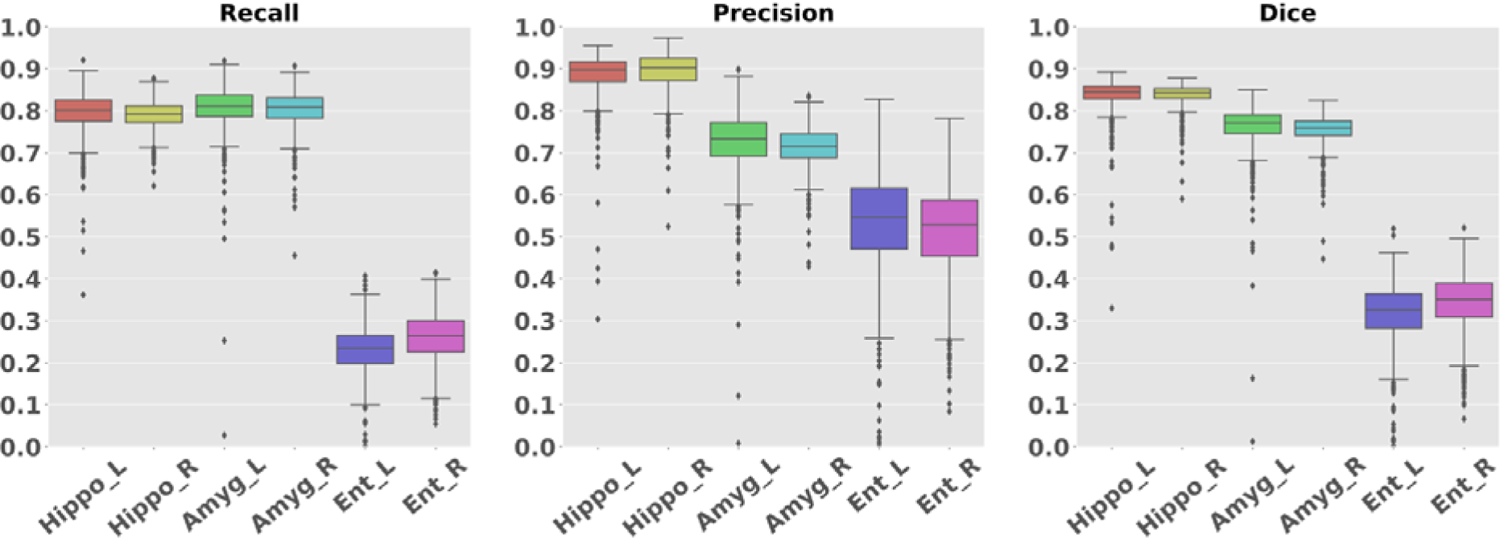
Boxplot of the Recall, Precision, and Dice scores in hippocampus, amygdala, and entorhinal regions.

**Figure 9.**
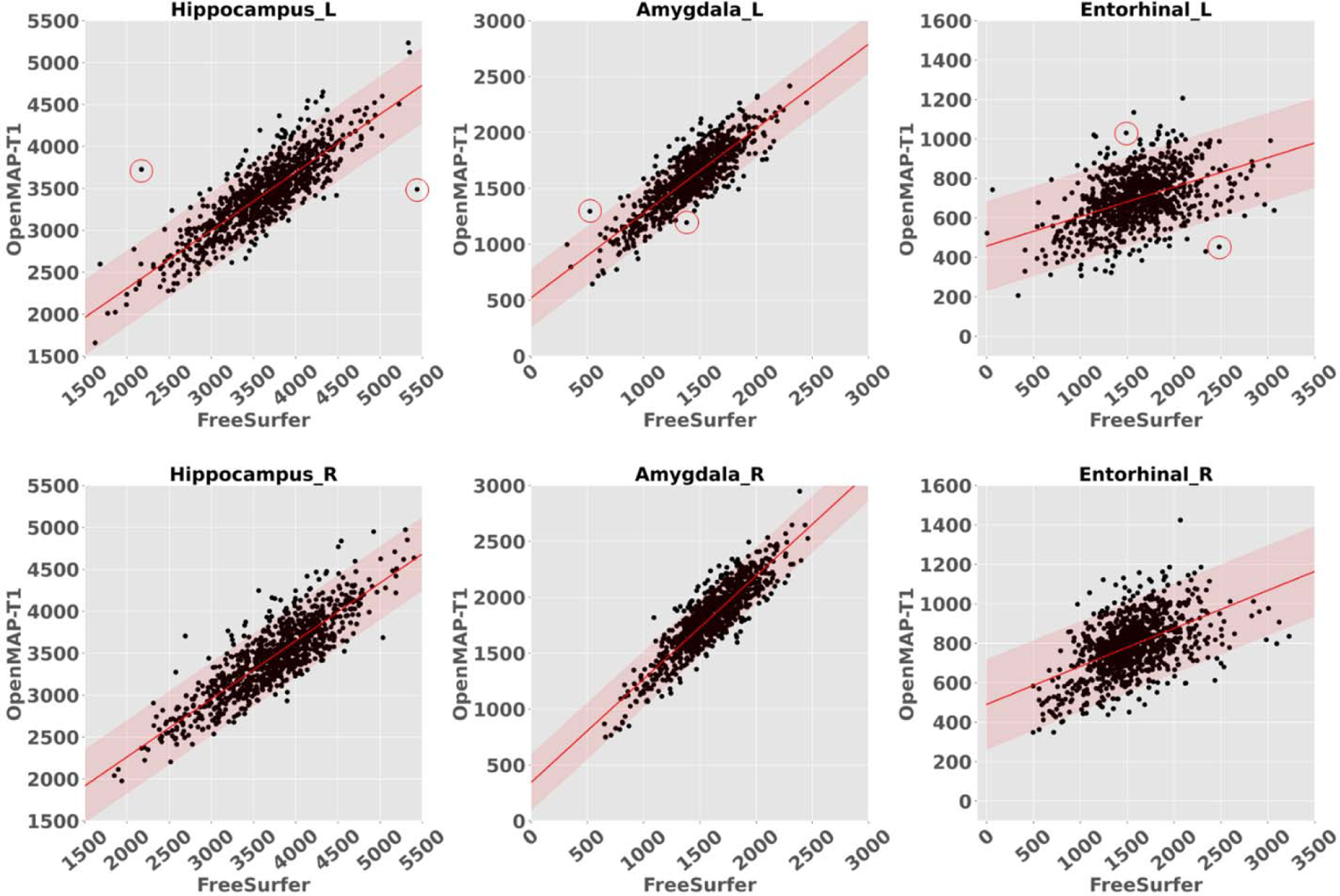
The correlations between the predicted volumes of the hippocampus, amygdala, and entorhinal cortex as obtained by OpenMAP-T1 and FreeSurfer. The upper row displays the results from the left side, while the lower row presents the results from the right side in ADNI3. The red lines represent the regression lines, while the transparent red areas show the range encompassing three standard deviations from these fitted regression lines. The red circles in the graph correspond to the representative images showcased in Figure 10.

**Figure 10.**
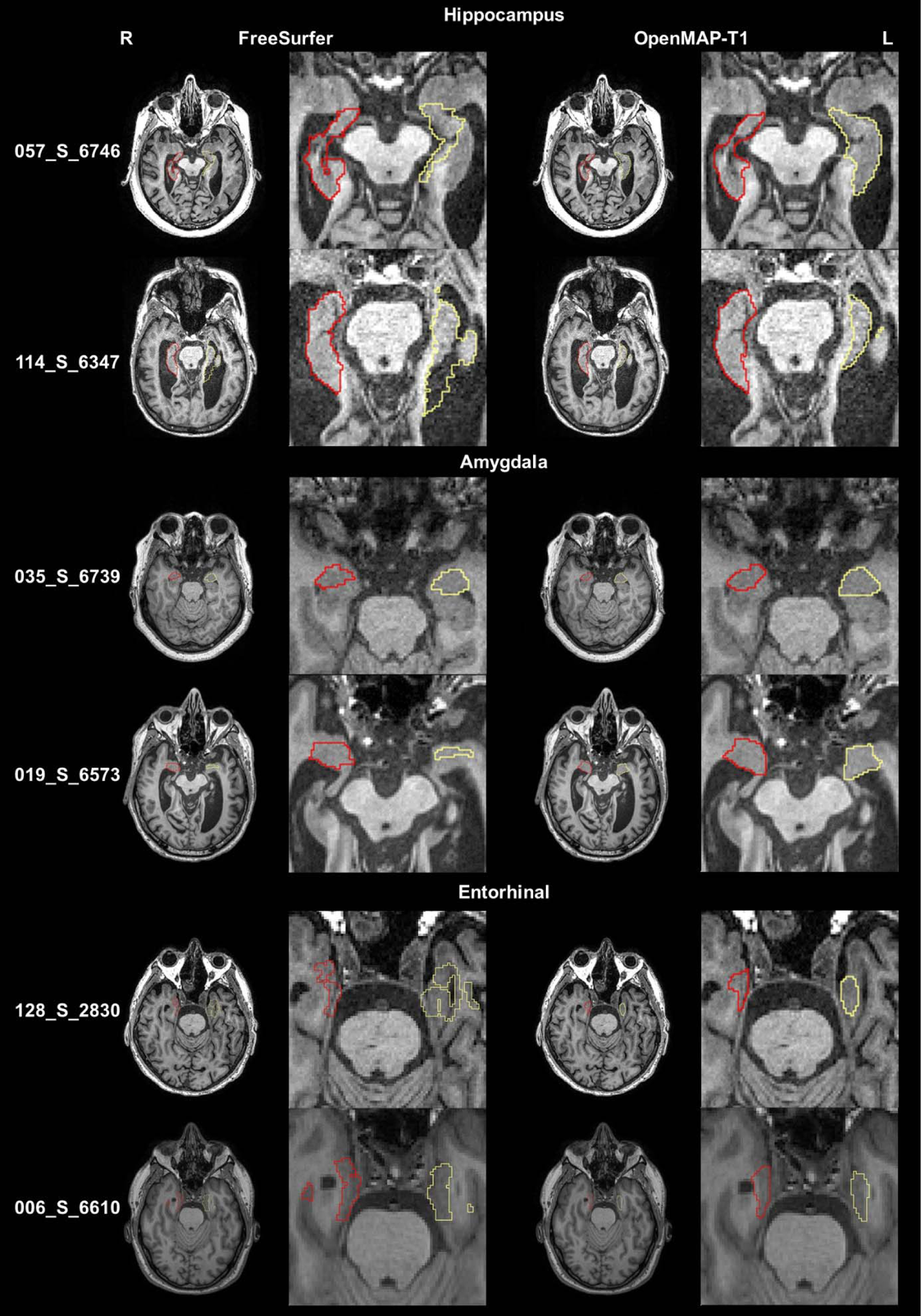
The parcellation maps from OpenMAP-T1 and FreeSurfer overlaid on the MPRAGE images that correspond to the red circles in Figure 9 are demonstrated. The leftmost column shows the study IDs from the ADNI3 dataset. Axial slices along with magnified images of the hippocampus, amygdala, and entorhinal cortex are presented. The red borderlines indicate the regions on the right side, while the yellow borderlines mark the regions on the left side. These images adhere to the radiological convention, where the left side of the brain is shown on the right side of the image.

Figure 10 displays representative images linked to the outliers in Figure 9. The primary reasons for the low Dice scores were generally due to mislabeling by FreeSurfer, differences in anatomical definitions between OpenMAP-T1 and FreeSurfer, or a combination of both. For instance, in image 057_S_6746, FreeSurfer incorrectly labeled the left hippocampus. In image 114_S_6347, FreeSurfer erroneously included a part of the lateral ventricle in the left hippocampus ROI. The boundary of the amygdala was delineated smaller by FreeSurfer than defined by OpenMAP-T1 in images 035_S_6739 and 019_S_6573, which is attributable to the differing anatomical definitions between the two. Furthermore, FreeSurfer typically incorporated the perirhinal cortex and the dura mater adjacent to the entorhinal cortex into the entorhinal ROI. In contrast, the definition of the entorhinal cortex in OpenMAP-T1 does not include these areas, as evident in images 128_S_2830 and 006_S_6610.

### 3.3. Biological Evaluation

AUC values for the ROC curves were 0.920 for MALF, 0.930 for OpenMAP-T1, and 0.908 for FreeSurfer. There were no significant differences between these AUC values. The p-values derived from the DeLong test were 0.618 when comparing MALF and OpenMAP-T1, and 0.271 when comparing OpenMAP-T1 and FreeSurfer.

Among the top 20 anatomical regions with the highest correlation coefficients derived from the trained LASSO model, nine structures were common between MALF and OpenMAP-T1 (Figure 11). Of these, six structures – namely the hippocampus, amygdala, inferior horn and body of the lateral ventricle, Sylvian fissure, and the hippocampal part of the cingulum bundle – are known to be associated with AD pathologies [65–68], indicating the appropriateness of the LASSO model.

**Figure 11.**
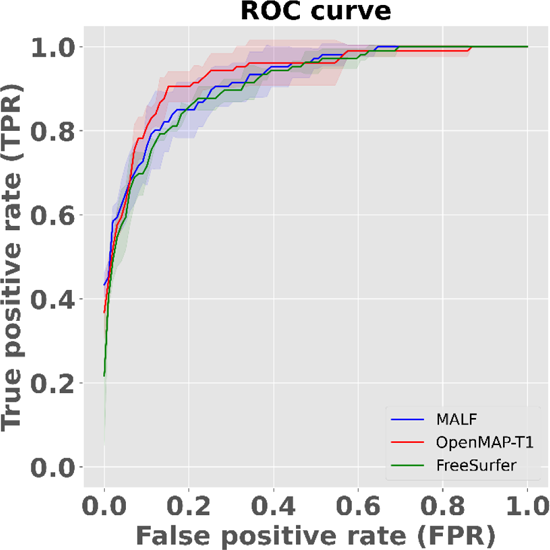
The Receiver Operatorating Characteristic (ROC) curve for distinguishing between AD and CN based on predicted volumes using the LASSO model. The blue line represents the ROC curve for MALF, the red line for OpenMAP-T1, and the green line for FreeSurfer. The shaded areas around each line indicate the standard deviation.

**Figure 12.**
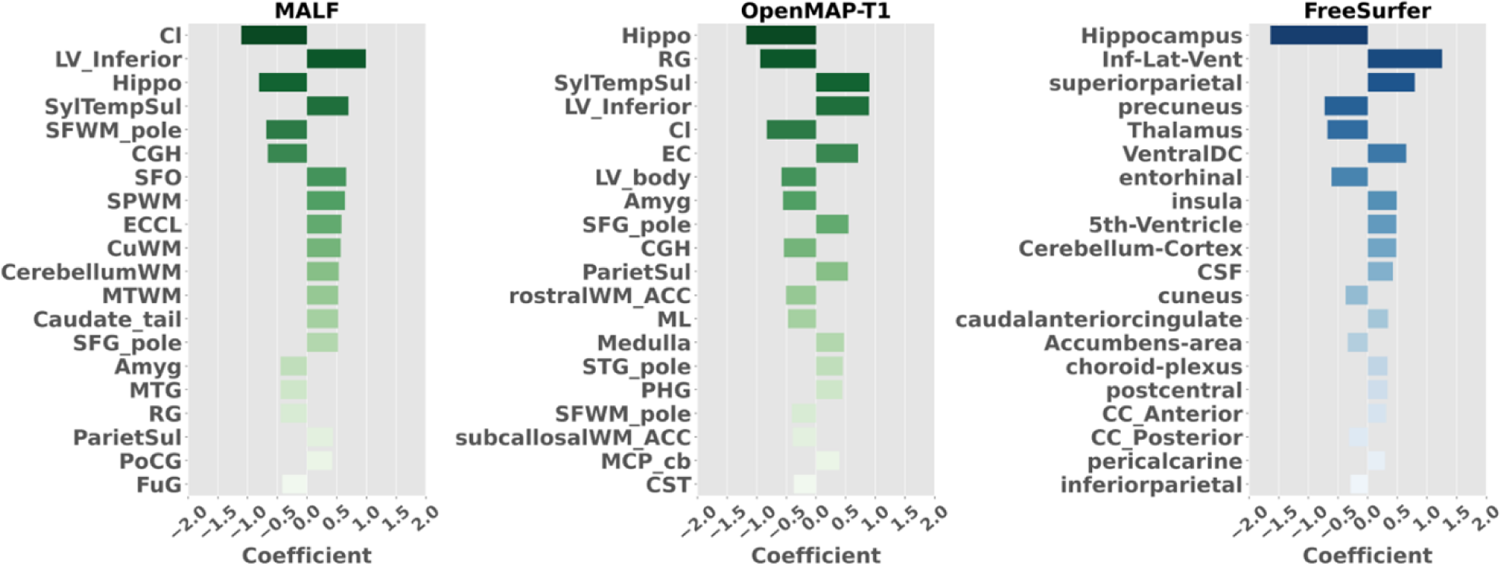
The 20 anatomical regions that exhibited the highest correlation coefficients as determined by the trained LASSO model.

## 4. Discussion

We have developed and released a deep-learning model, called OpenMAP-T1, on Git-Hub, for the segmentation and parcellation of brain 3D T1-weighted MRI images, based on their anatomical structures. The performance of the model was evaluated using metrics such as recall, precision, Dice, and correlation based on comparisons between the OpenMAP-T1 and the established MALF algorithm. The results demonstrated that the parcellation outcomes were generally equivalent to the MALF method. Notably, the processing time was significantly reduced to less than 90 seconds per image, compared to the several hours required by the existing MALF method.

We introduced a multi-processing phase to create a parcellation model, which is robust against diverse imaging environments. In MRI parcellation using deep-learning, performance is often affected by differences in the imaging environment, particularly the defacing algorithm, between training and evaluation datasets [69]. To reduce the impact of irrelevant areas on whole-brain parcellation, we incorporated cropping and skull-stripping phases. Furthermore, the whole brain was parcellated into 141 regions combining left and right labels, inspired by the approach used in FastSurfer [44]. While FastSurfer combines the left and right labels using only the sagittal cross-section, OpenMAP-T1 can combine the left and right labels in all the cross-sections. Consequently, we could reduce the number of class (i.e., region labels) and increase the size of each region, which resulted in fast and accurate parcellation.

There is the potential to further enhance the processing speed of OpenMAP-T1. For instance, reducing the image matrix size could decrease the input size for the PNet. This reduction can be achieved by repositioning the center of gravity of the brain post skull-stripping, and then trimming areas outside the brain. We anticipate periodic updates to OpenMAP-T1, focusing on optimizing processing strategies and incorporating new features into the algorithms.

In recent years, the MPRAGE sequence with 3T scanners has been often used for anatomical MRI scans for brain research [70]. However, available open brain MRI databases contain legacy images scanned with MRI scanners from various vendors, models, and magnetic field strengths. These databases also include images scanned with the MPRAGE and other sequences, such as the SPGR sequence. The accuracy of brain parcellation was affected by variations in scanner types, magnetic field strengths, and scan protocols [71, 72]. Thus, automated brain MRI parcellation methods should be robust to these variations. Furthermore, it has been demonstrated that facial appearance can be reconstructed from a whole-head MRI [73]. In terms of a privacy protection perspective, some MRI databases [44] apply a defacing algorithm to prevent facial reconstruction. The ability of automated brain parcellation methods to handle such defaced MRIs is crucial in the era of open science and data sharing. Our results showed that the parcellation maps generated by MALF and OpenMAP-T1 are substantially similar (average Dice score > 0.8, except for LBPA40 database that collected SPGR images) regardless of the scanner vendor, magnetic field strength, scan protocol, or the presence of defacing. However, we found that Dice scores were significantly lower in SPGR images than those in MPRAGE images, from 1.5T to 3T scanners, as well as the Dice scores in GE scanners, which were significantly lower compared to those in Philips or Siemens.

We identified three major reasons that MALF and OpenMAP-T1 showed disagreement (Dice score < 0.8). MALF tended to mislabel images with noticeable intensity inhomogeneity even after correction with the N4 algorithm. For instance, images scanned with 3D-SPGR sequences or 1.5T scanners resulted in blurring contrasts between the white matter and gray matter compared to MPRAGE or 3T scanners, or images that had undergone defacing resulted in low Dice scores as well. These low Dice scores were not caused by mislabeling by OpenMAP-T1, but rather, by MALF. MALF uses whole-head MPRAGE images scanned with 3T scanners as training data and employs image transformation using intensity information as a cost function [38]. Therefore, MALF is vulnerable to intensity inhomogeneity, and its parcellation accuracy might be affected by differences in scanner magnetic field strength, scan protocols, and defacing. In contrast, OpenMAP-T1, which uses deep-learning with data augmentation, was more robust than MALF against variations in image intensity inhomogeneity and scanning parameters.

Images with notable intensity inhomogeneity, even after intensity correction, were often scanned in a head-extended position. Such position is often seen in older participants with neurological conditions [74, 75], leading to a decrease in image intensity in areas distant from the physical center of the MRI scanner, such as the frontal pole or the posteroinferior occipital lobe and cerebellum. The robustness of OpenMAP-T1 for the abnormal head position seems advantageous for the analysis of disease images. Moreover, artifacts that introduce high-signal pixels potentially hinder successful intensity correction. The results with OpenMAP-T1 indicated its robustness against such artifacts.

Defacing may become a standard procedure when sharing brain MRI images [76–78]. However, many brain parcellation methods have not been tested on images that have undergone defacing. Implementing a refacing process, which adds artificial facial information after defacing, is being considered to avoid changes in parcellation accuracy due to defacing [73]. Meanwhile, OpenMAP-T1 has demonstrated its capability to parcellate images from the NFBS and OASIS1 databases, which have used different defacing methods, indicating its adaptability for future mainstream databases of defaced images.

From the perspective of precision medicine, a significant topic has been whether insights gained from basic research and clinical trials can be adapted to actual clinical data. Research often involves strict inclusion and exclusion criteria, targeting specific individuals, inevitably leading to selection bias [79–81]. Therefore, using real-world data is essential when considering the applicability to clinical data. For developing automated brain MRI parcellation methods, assessing their accuracy using brain images obtained through clinical practice is crucial. To evaluate the applicability of OpenMAP-T1 in real clinical settings, we used real-world data from OASIS4. The average Dice score exceeded 0.8 in OASIS4, suggesting potential adaptability to images obtained through clinical practice.

When testing research data such as ADNI, the Dice score was unaffected by sex or diagnostic category, including cognitively normal (CN), MCI, and AD. However, in the real-world data of OASIS4, while the Dice score was not influenced by age or sex, it was affected by clinical diagnosis. Average Dice scores of groups with “other non-AD neurodegenerative disorders” and “vascular cognitive impairment” tended to be lower than other diagnostic groups, although they still demonstrated average Dice scores above 0.8. Many images with Dice scores below 0.8 were due to mislabeling by MALF in the presence of lesions, but, in patients with large arachnoid cysts at the posterior area, both MALF and OpenMAP-T1 mislabeled the lesion. This result suggests that OpenMAP-T1 may mislabel images containing large lesions, likely due to a lack of such images in its training data, indicating potential areas for future improvement.

FreeSurfer is one of the most commonly used softwares for brain MRI parcellation [82, 83]. FreeSurfer is suitable for comprehensive analysis of the cerebral cortex with its ability to measure the thickness of the cerebral cortex. However, OpenMAP-T1 can parcellate both gray and white matter areas, thus offering the advantage of simultaneous analysis of the cortex and white matter. Since the definitions of the brain regions employed by both softwares differ, and their intended uses are different, it is difficult to compare the parcellation performance of the two. Therefore, their performance was compared in a task that separates brain MRIs of AD and CN individuals. In AD research, the amygdala, entorhinal cortex, and hippocampus volumes are often used as neurodegeneration markers. When comparing these volumes measured by FreeSurfer and OpenMAP-T1, a good correlation was found in these regions. Furthermore, the brain regions that played a significant role in separating AD and CN, obtained from the LASSO model, were similar, and there was no difference between FreeSurfer and OpenMAP-T1 in separation performance for AD and CN as analyzed by ROC analysis. Therefore, the choice between FreeSurfer and OpenMAP-T1 should not be based on which has more accurate parcellation, but should be selected according to the research objectives and the anatomical structures of interest.

## 5. Limitations

OpenMAP-T1 has several limitations. As with any parcellation method, in small ROIs, precision, recall, and Dice scores can be significantly reduced by minor boundary differences, making it challenging to evaluate the accuracy of parcellation itself. Furthermore, whether parcellation can be accurately performed on images containing various lesions, such as large cerebral infarcts, brain hemorrhages, or brain tumors, is a topic for future investigation. Using real clinical images as training data for clinical applications might be necessary. Also, while the analyzed images in this study were high-resolution 3D images, many clinical images use 2D imaging methods with a thickness of 5mm or more. Whether OpenMAP-T1 can be applied to such thick-slice 2D images is also a subject for future study.

## 6. Conclusion

We developed OpenMAP-T1, a model based on deep-learning for segmentation and parcellation of brain T1-weighted MRI images according to anatomical structures, and evaluated its accuracy across eight test datasets. OpenMAP-T1 could accurately perform parcellation regardless of technological variations such as the scanner vendor, magnetic field strength, defacing, imaging parameters, defacing, and biological variations, including differences in sex, age, and disease. It could also parcellate images with postural changes in the head or images with intensity inhomogeneity. In a task using machine-learning to differentiate between AD and CN based on parcellation maps, the discriminative ability of OpenMAP-T1 was found to be equivalent to that obtained using MALF or FreeSurfer. However, OpenMAP-T1 was able to process images much faster compared to the traditional MALF method and FreeSurfer. These results suggest that OpenMAP-T1 is a promising method for high-speed image parcellation that can accommodate various types of lesions. OpenMAP-T1 is available on our GitHub (URL: https://github.com/OishiLab/OpenMAP).

## Data Availability

All data produced are available online at: https://github.com/OishiLab/OpenMAP-T1-V2

## Acknowledgment

The MRI data collection and sharing for this project was funded by the Alzheimer’s Disease Neuroimaging Initiative (ADNI) (National Institutes of Health Grant U01 AG024904) and DOD ADNI (Department of Defense award number W81XWH-12-2-0012). ADNI is funded by the National Institute on Aging, the National Institute of Biomedical Imaging and Bioengineering, and through generous contributions from the following: AbbVie, Alzheimer’s Association; Alzheimer’s Drug Discovery Foundation; Araclon Biotech; BioClinica, Inc.; Biogen; Bristol-Myers Squibb Company; CereSpir, Inc.; Cogstate; Eisai Inc.; Elan Pharmaceuticals, Inc.; Eli Lilly and Company; EuroImmun; F. Hoffmann-La Roche Ltd. and its affiliated company Genentech, Inc.; Fujirebio; GE Healthcare; IXICO Ltd.; Janssen Alzheimer Immunotherapy Research & Development, LLC.; Johnson \& Johnson Pharmaceutical Research & Development LLC.; Lumosity; Lundbeck; Merck & Co., Inc.; Meso Scale Diagnostics, LLC.; NeuroRx Research; Neurotrack Technologies; Novartis Pharmaceuticals Corporation; Pfizer Inc.; Piramal Imaging; Servier; Takeda Pharmaceutical Company; and Transition Therapeutics. The Canadian Institutes of Health Research is providing funds to support ADNI clinical sites in Canada. Private sector contributions are facilitated by the Foundation for the National Institutes of Health (www.fnih.org). The grantee organization is the Northern California Institute for Research and Education, and the study is coordinated by the Alzheimer’s Therapeutic Research Institute at the University of Southern California. ADNI data are disseminated by the Laboratory for Neuro Imaging at the University of Southern California.

Data were provided by OASIS1/4: Cross-Sectional: Principal Investigators: D. Marcus, R. Buckner, J. Csernansky, J. Morris; P50 AG05681, P01 AG03991, P01 AG026276, R01 AG021910, P20 MH071616, U24 RR021382 and Clinical Cohort: Principal Investigators: T. Benzinger, L. Koenig, P. LaMontagne. We would like to thank Jill Chotiyanonta for providing feedback on the accuracy of brain MRI parcellation results obtained from MRICloud, and Mary McAllister for her assistance with editing the English.

## SUPPLEMENTARY MATERIALS

**Supplementary Figure A.**
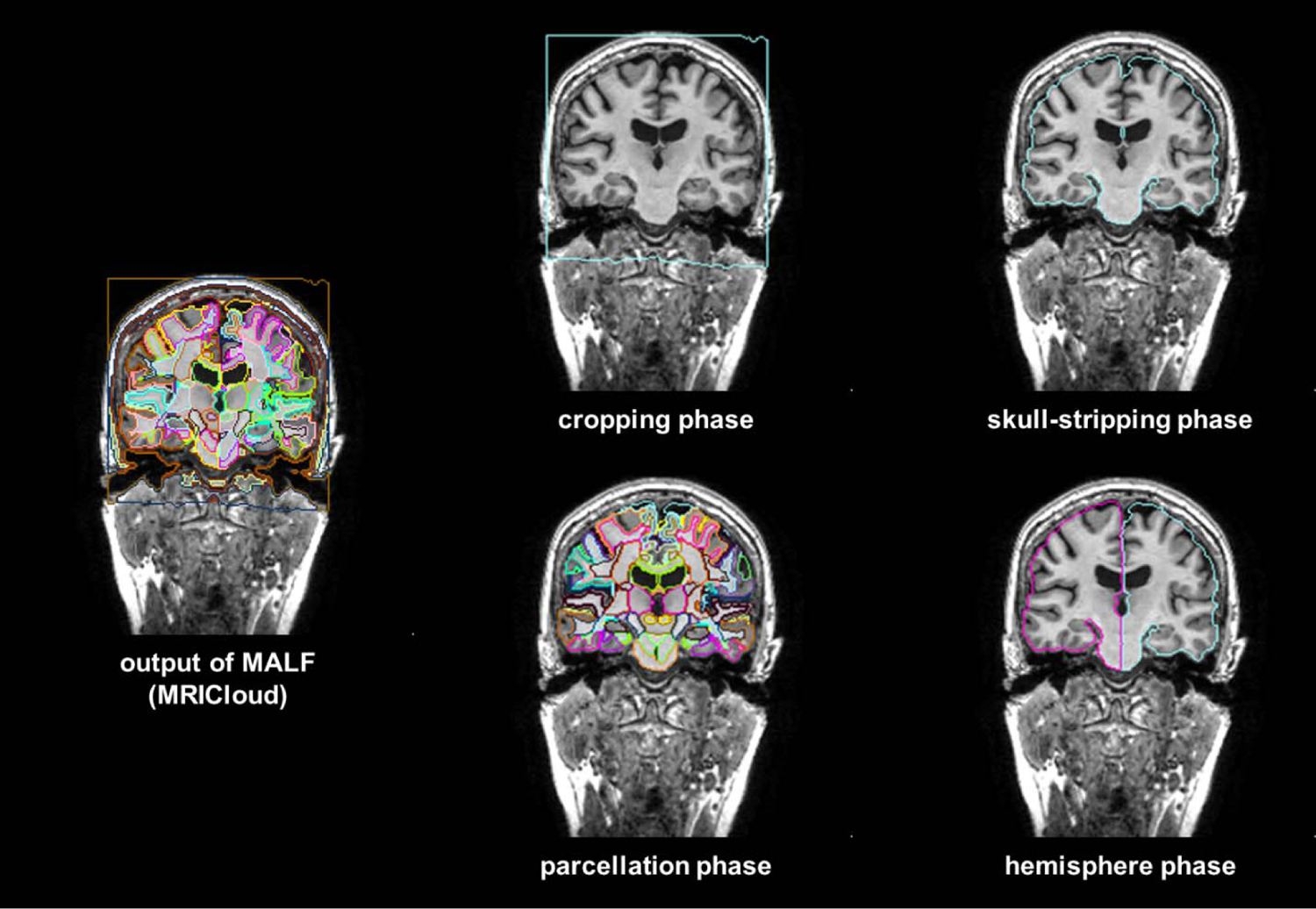
The procedure used to create training labels. The training labels for OpenMAP-T1 were generated by combining parcellation maps created by MALF (MRICloud).

**Supplementary Figure B.**
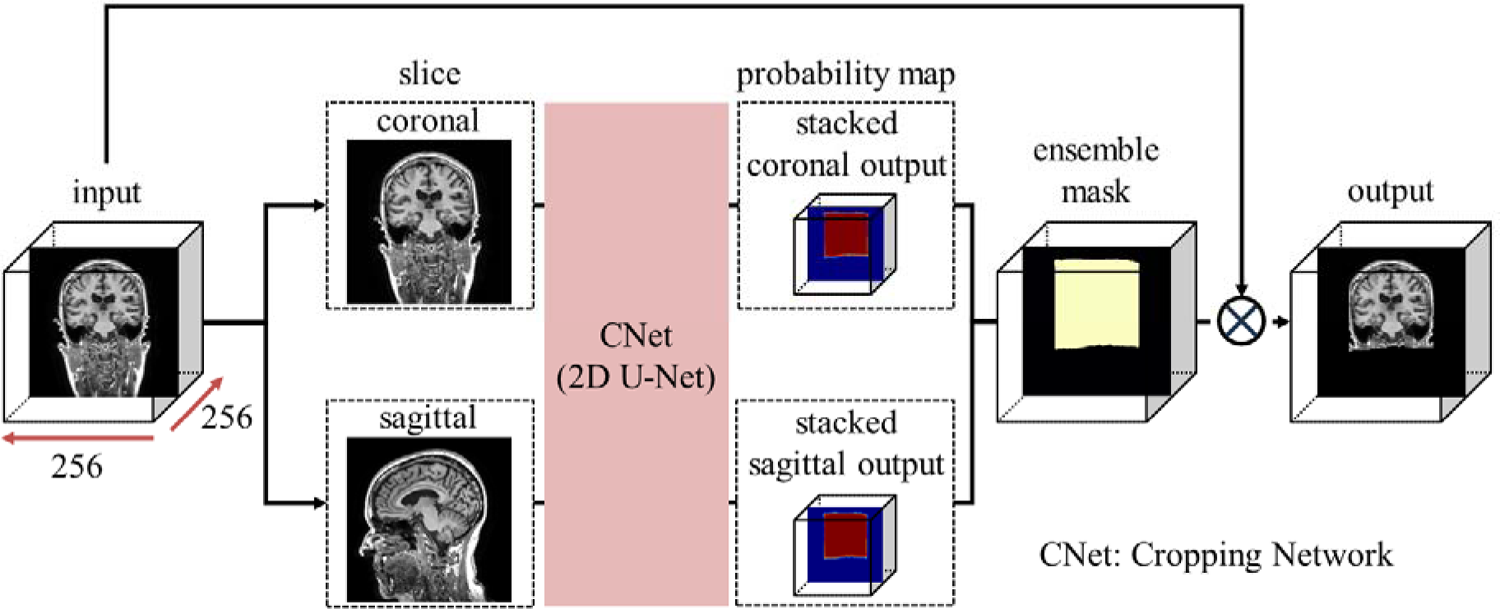
Overview of the cropping phase.

**Supplementary Figure C.**
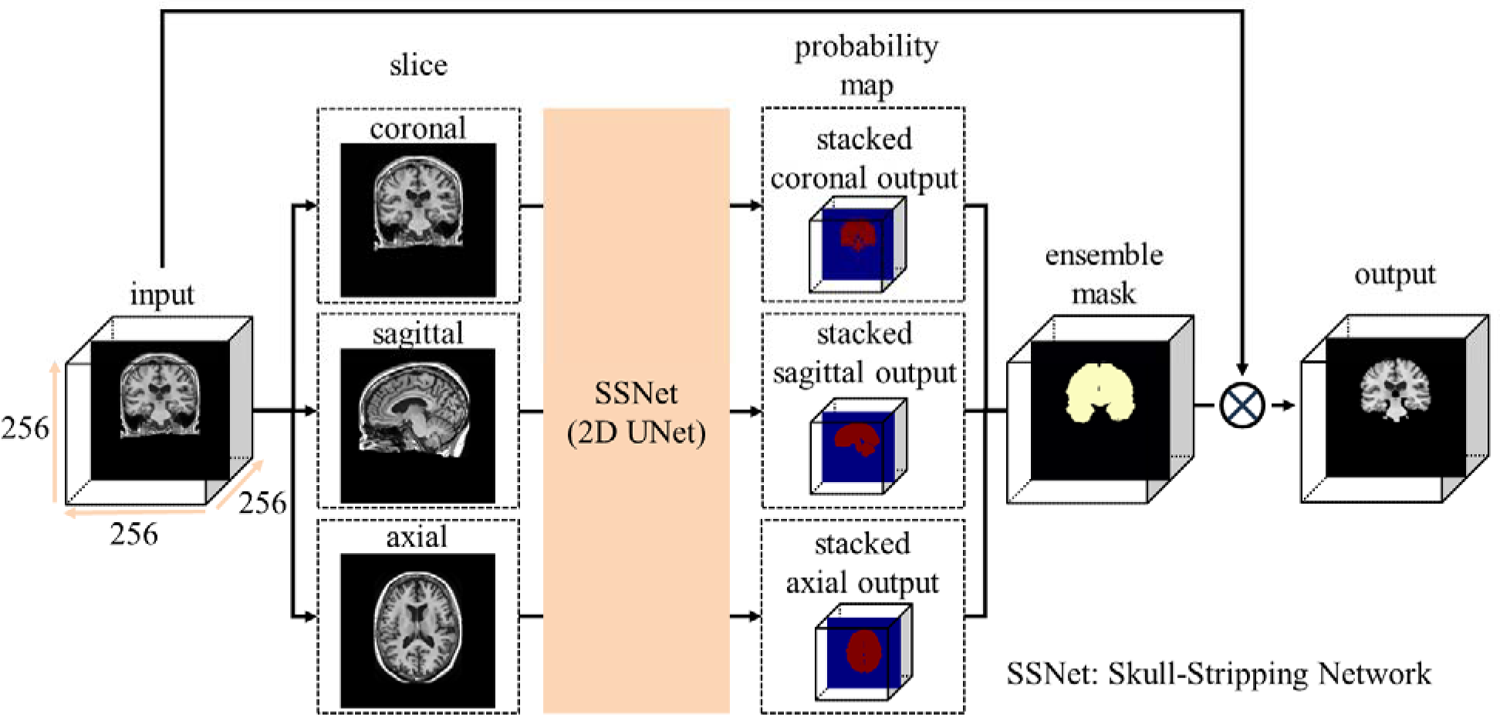
Overview of the skull-stripping phase.

**Supplementary Figure D.**
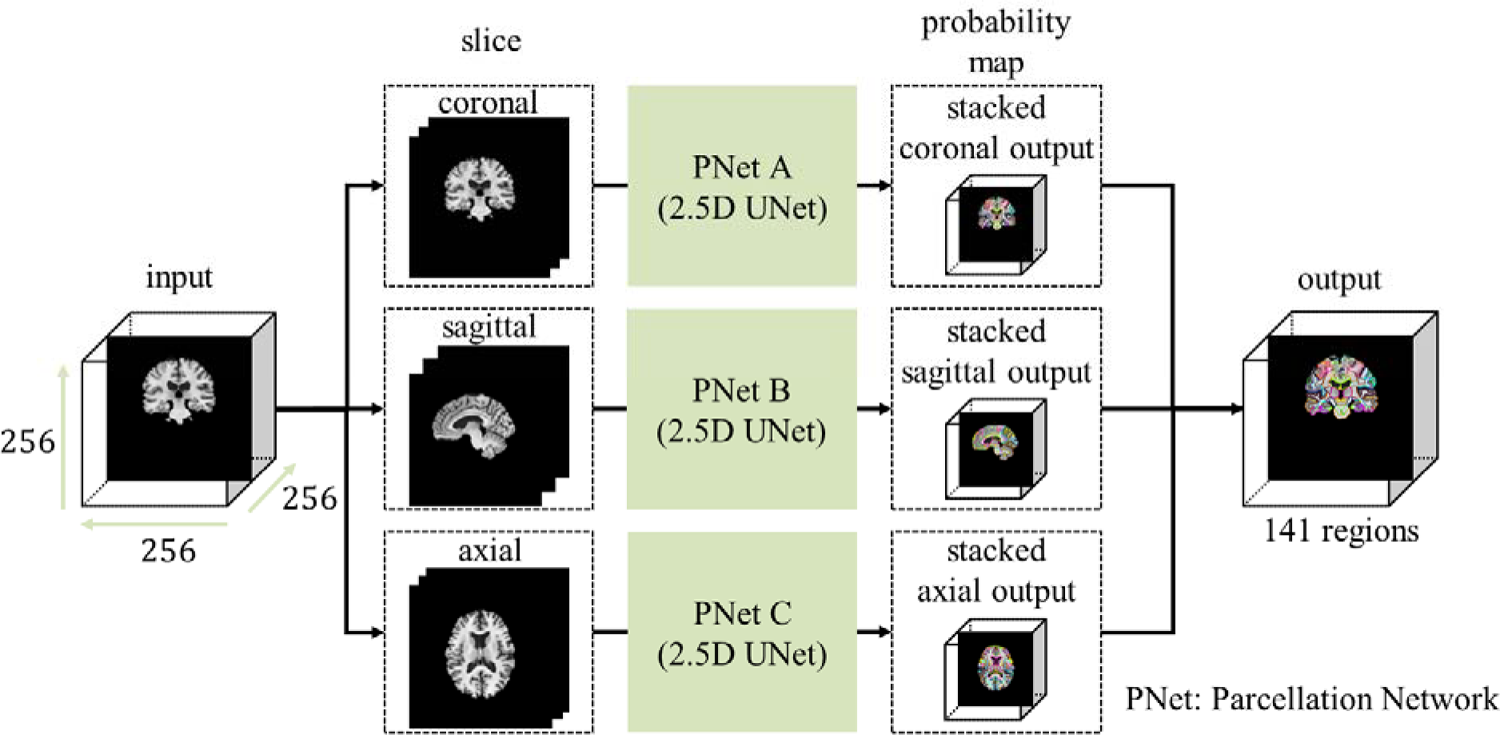
Overview of the parcellation phase.

**Supplementary Figure E.**
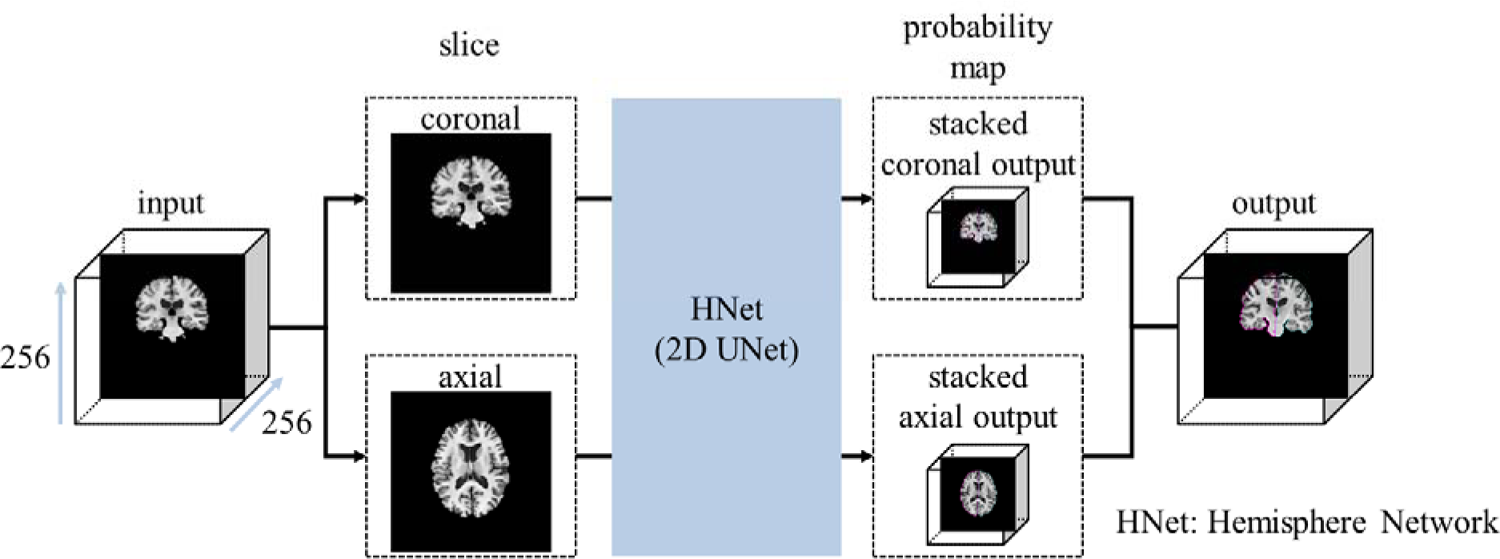
Overview of the hemisphere phase.

**Supplementary Figure F.**
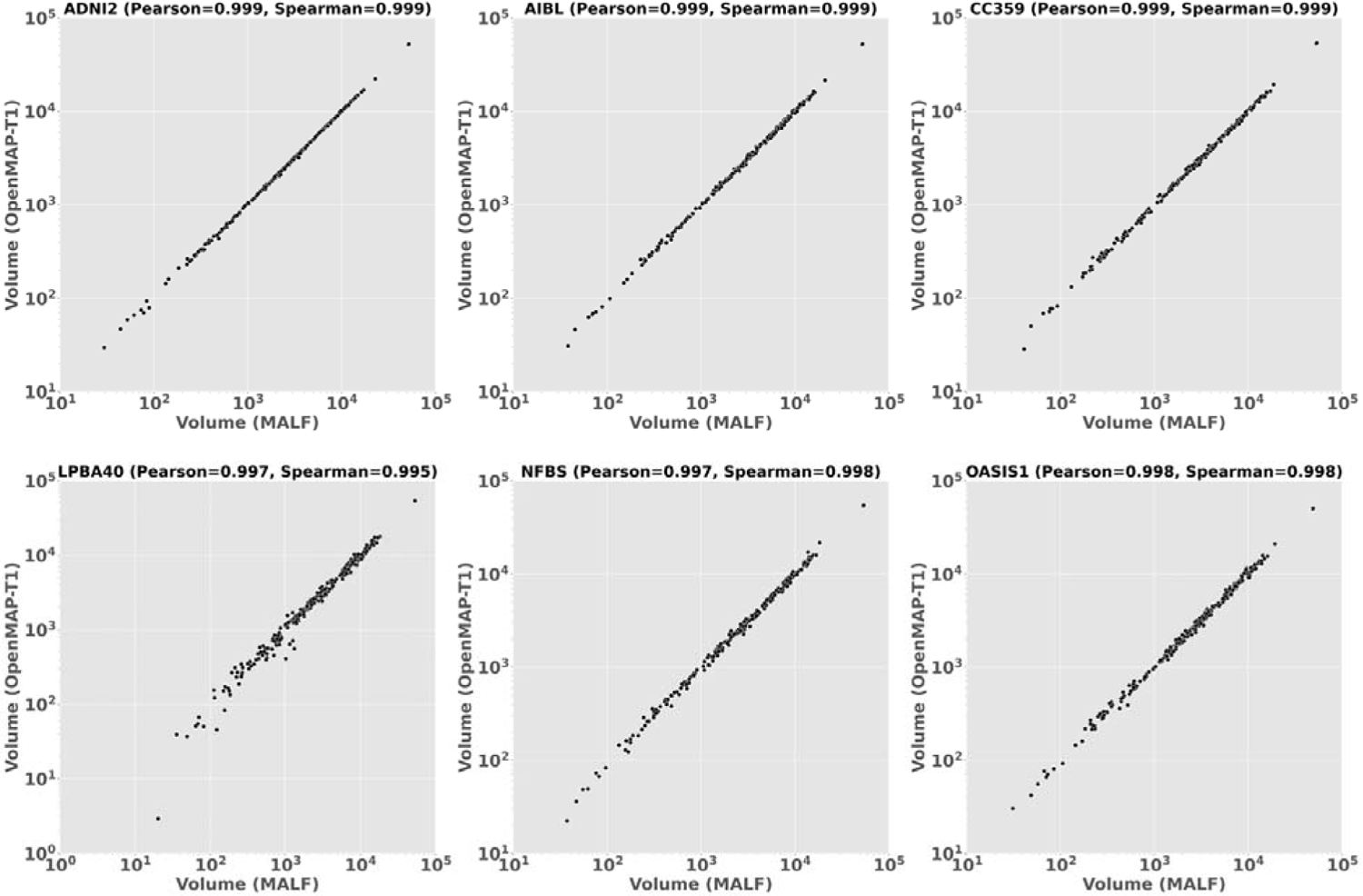
Correlation between the predicted volumes obtained using MALF and OpenMAP-T1 in ADNI2, AIBL, CC359, LPBA40, NFBS, and OASIS1. Note that only the LPBA40 has different scales for the x and y axes.

**Supplementary Figure G.**
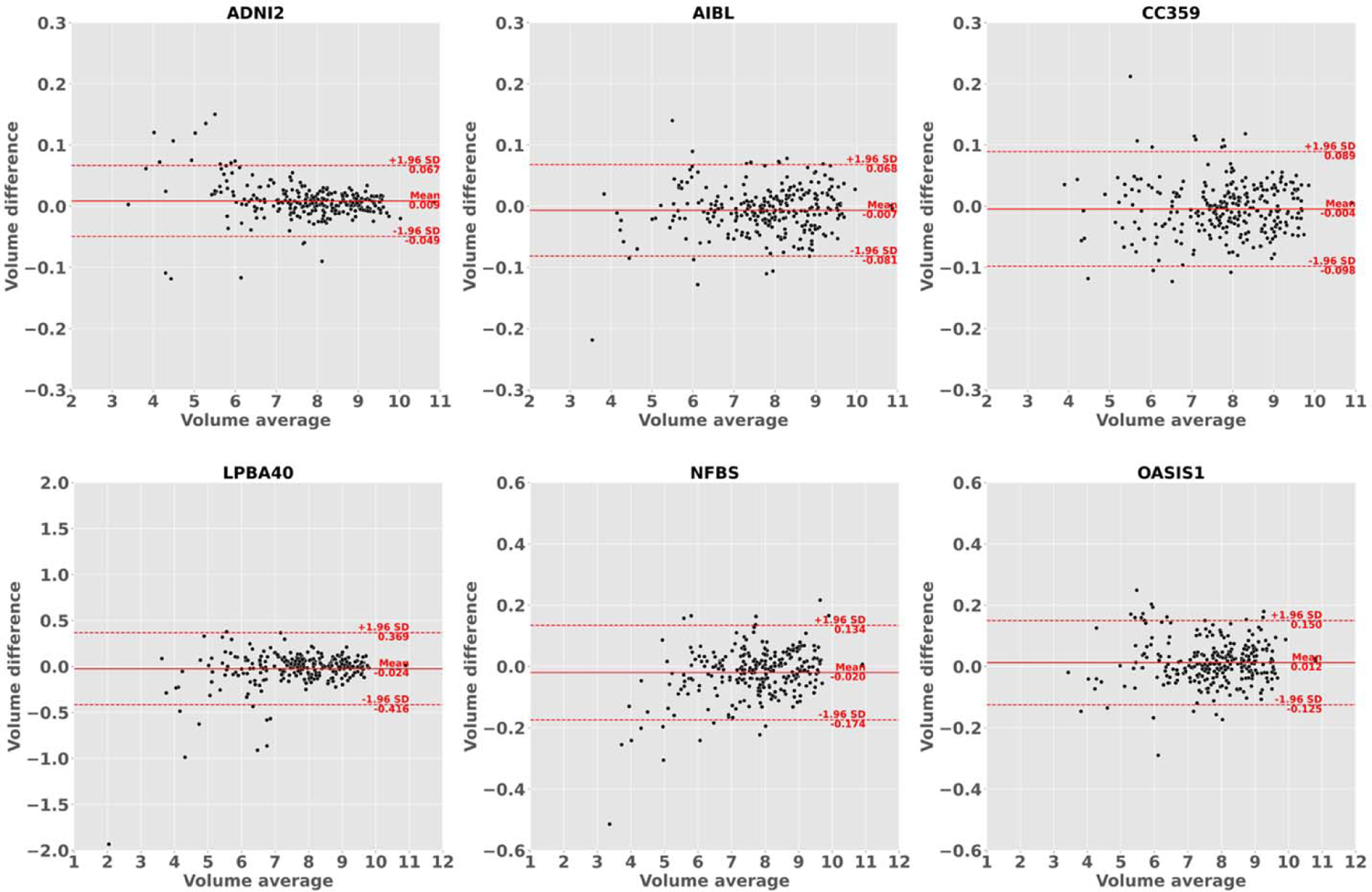
Bland-Altman plot demonstrates agreement between regional volumes predicted by MALF and OpenMAP-T1 in ADNI2, AIBL, CC359, LPBA40, NFBS, and OASIS1. The volume measurements were transformed using a base-2 logarithmic scale. Note that only the LPBA40, NFBS, and OASIS1 have different scales for the x and y axes.

**Supplementary Figure H.**
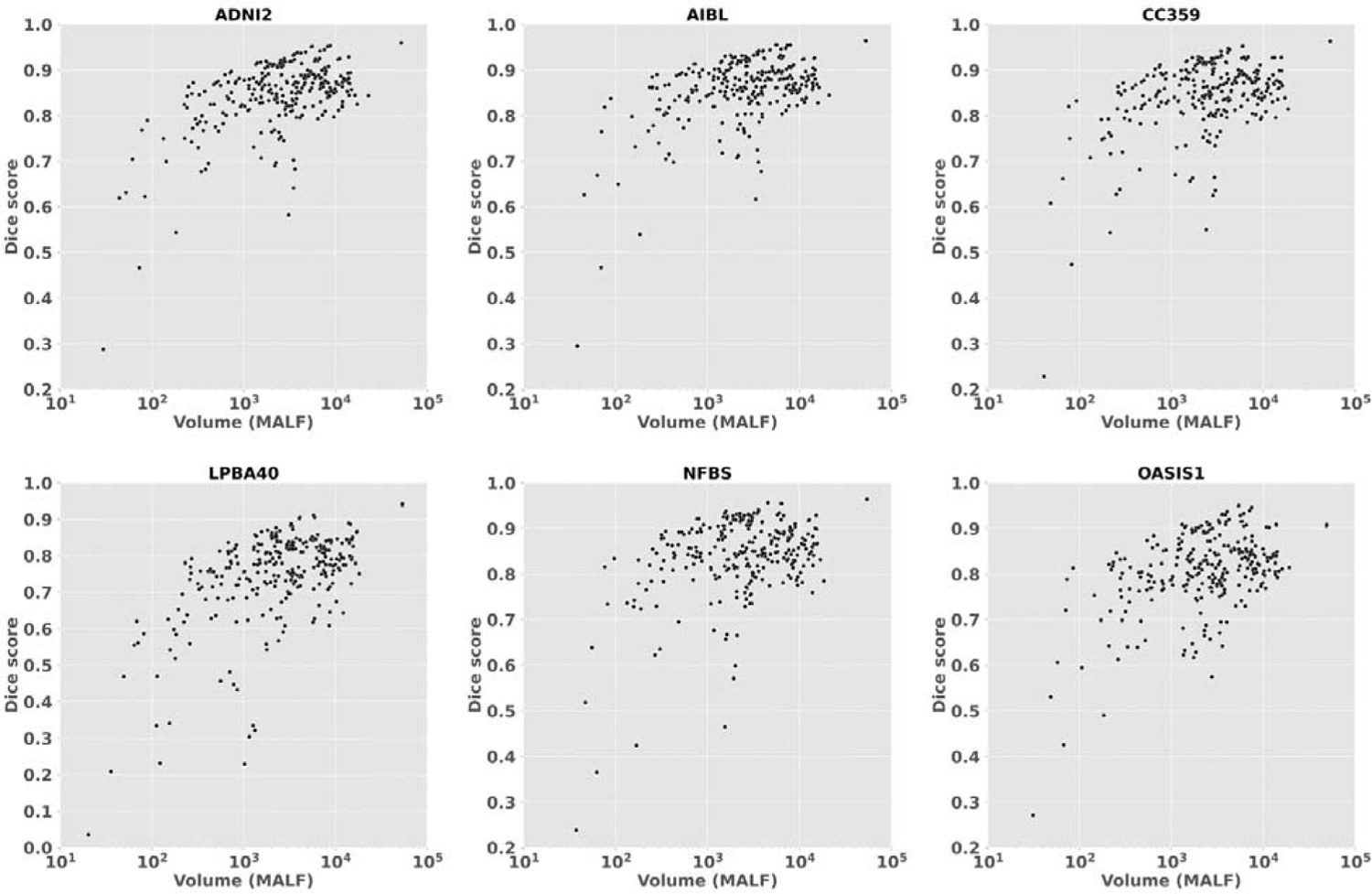
Relationship between the predicted volumes by MALF and Dice score in ADNI2, AIBL, CC359, LPBA40, NFBS, and OASIS1. Note that only the LPBA40 has different scales for the y axes.

---

**Table A1.**
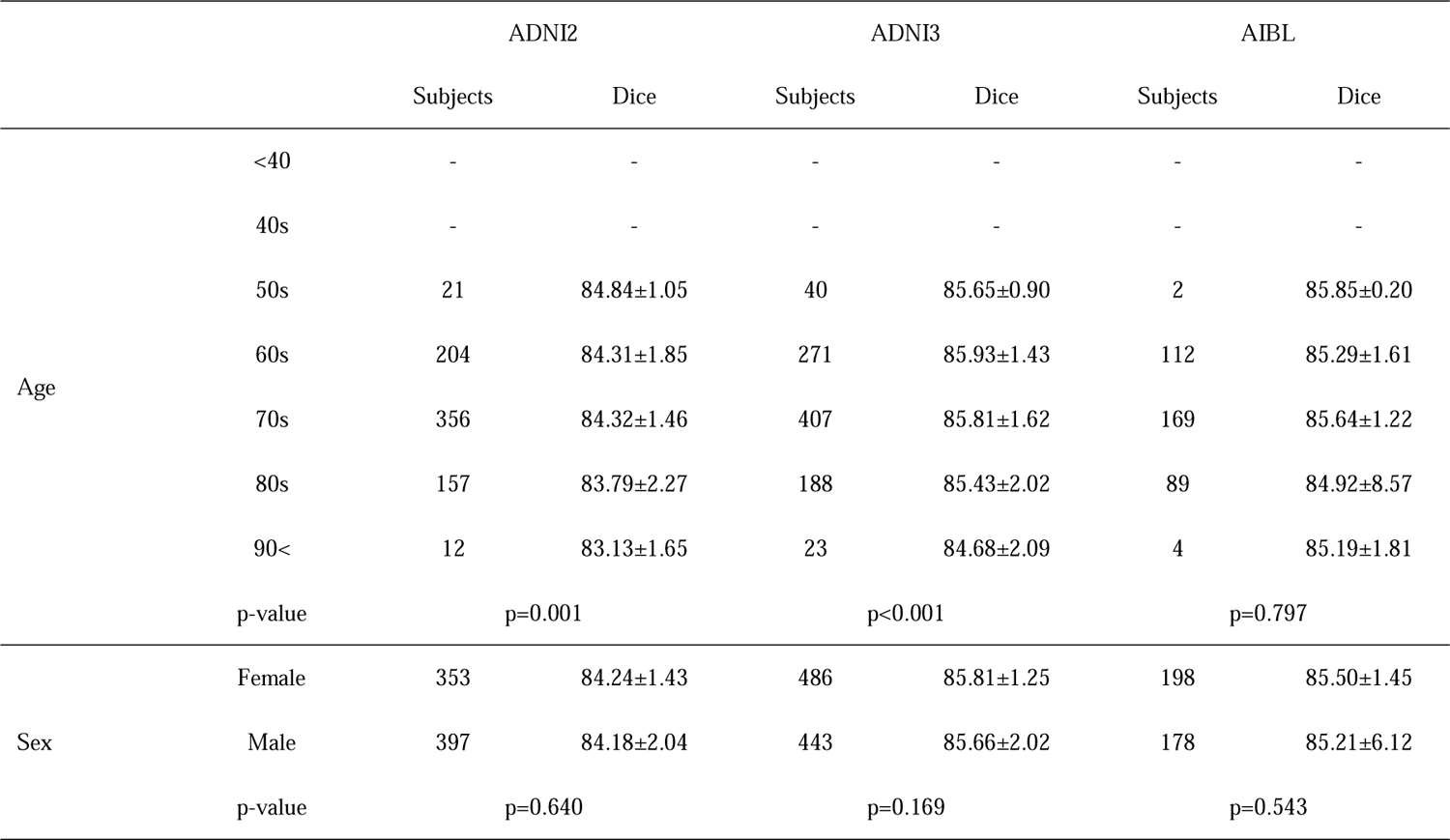
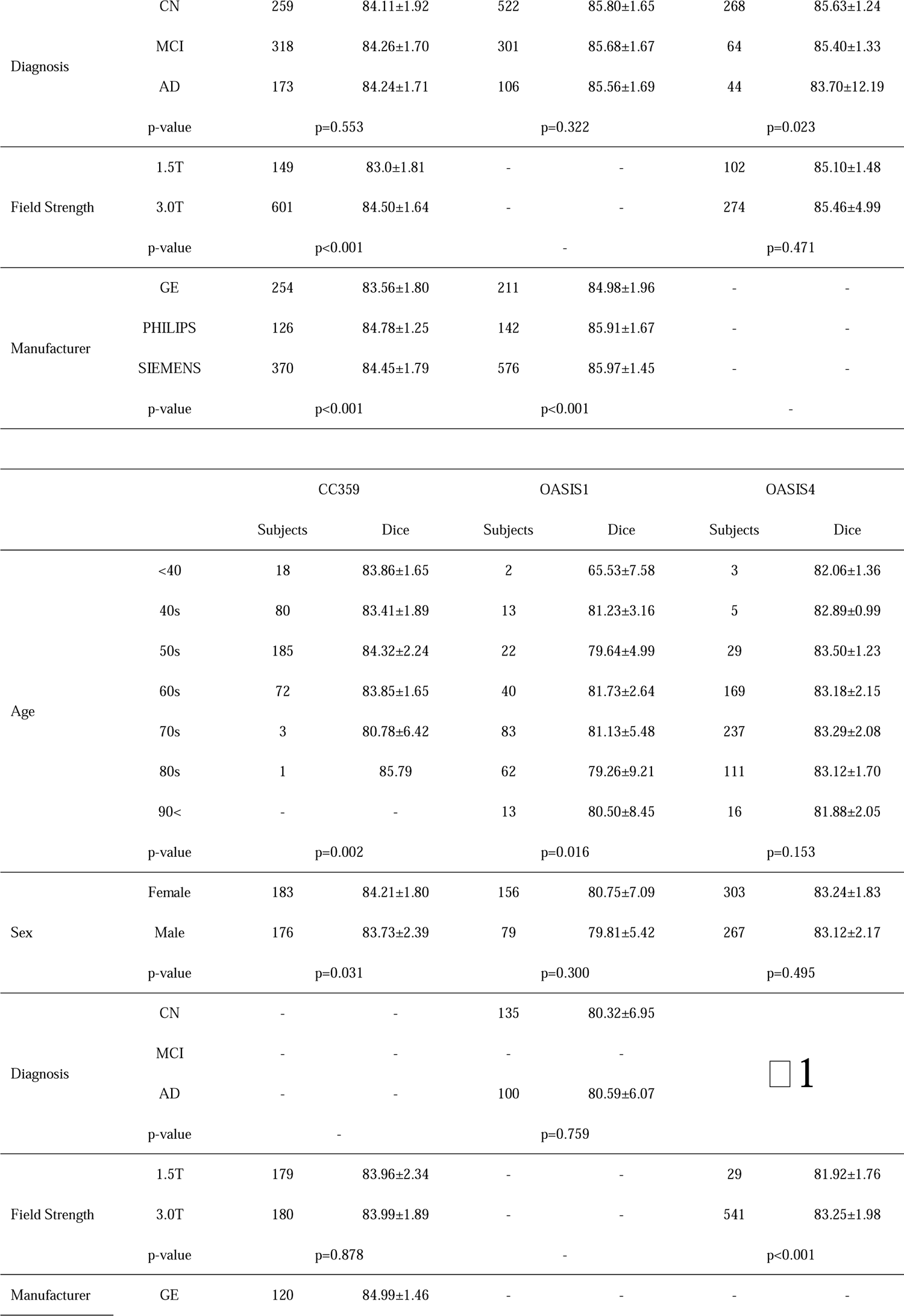
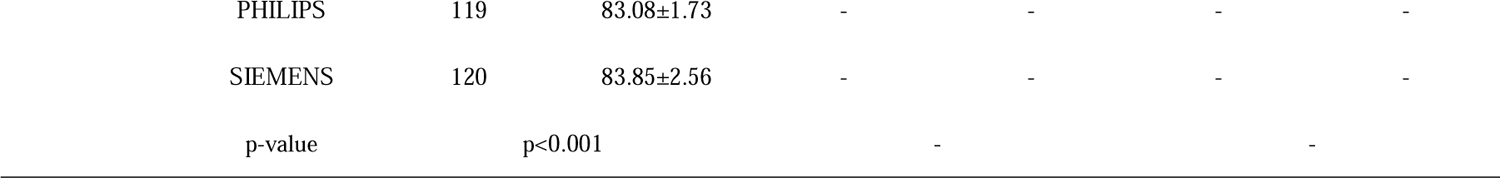
Dice score and P value (ANOVA) for each effect in all datasets.

**Table A2.** Dice score and p value (ANOVA) for diagnosis in OASIS4.

|  |  | OASIS4 |  |
| --- | --- | --- | --- |
|  |  | Subjects | Dice |
| ✖1<br>diagnosis | AD variant | 10 | 83.85±1.08 |
|  | AD + non neurodegenerative | 8 | 84.40±0.89 |
|  | AD / vascular | 38 | 82.89±2.01 |
|  | AD dementia | 189 | 83.26±1.88 |
|  | CN | 39 | 83.38±1.71 |
|  | Dementia with Lewy Bodies (DLB) | 11 | 83.34±1.83 |
|  | early onset AD | 23 | 83.23±2.35 |
|  | Frontotemporal Dementia (FTD) | 15 | 82.27±2.53 |
|  | Mild Cognitive Impairment (MCI) | 41 | 83.24±1.73 |
|  | mood / polypharmacy / sleep | 29 | 83.26±1.69 |
|  | non-neurodegenerative neurologic disease | 19 | 82.45±1.96 |
|  | other - miscellaneous | 15 | 83.00±1.84 |
|  | other non-AD neurodegenerative disorder | 7 | 81.17±4.08 |
|  | Primary Progressive Aphasia (PPA) | 7 | 84.00±1.04 |
|  | uncertain - AD possible | 105 | 83.46±2.07 |
|  | vascular cognitive impairment (VCI) | 14 | 81.36±2.47 |
| p-value |  | p=0.003 |  |

**Supplementary Table B.**
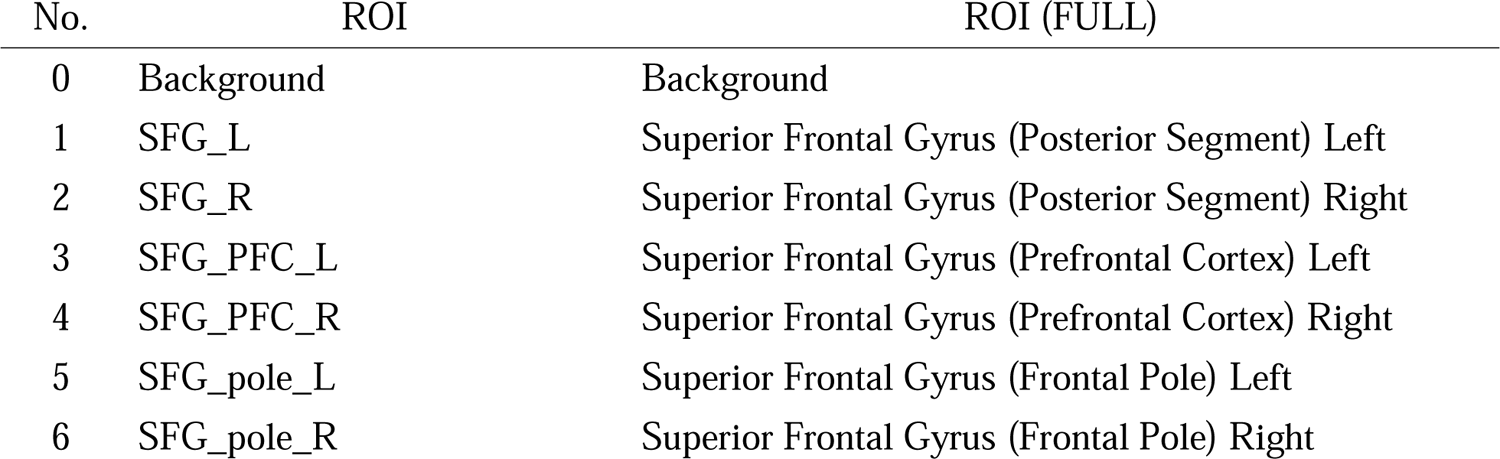
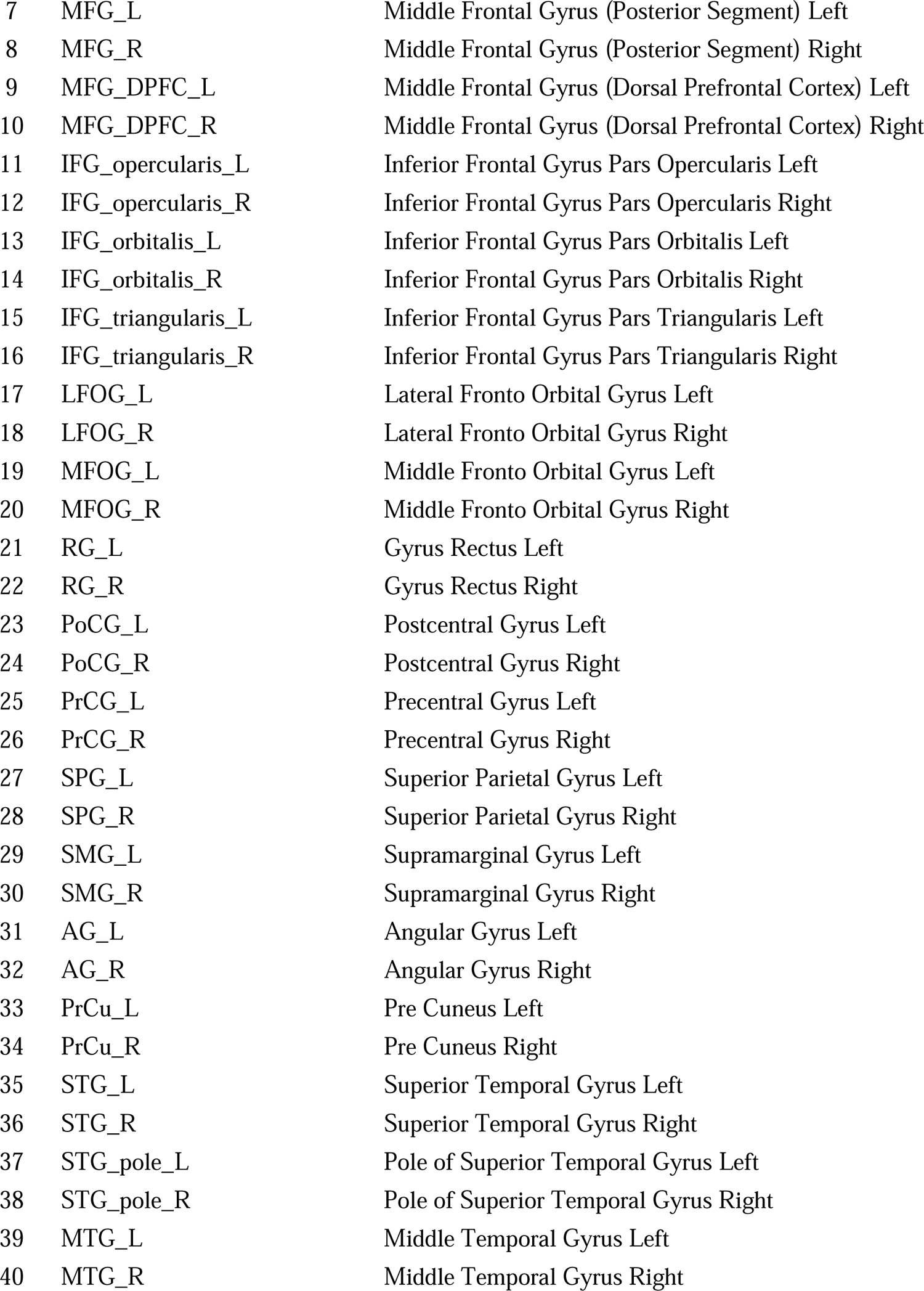
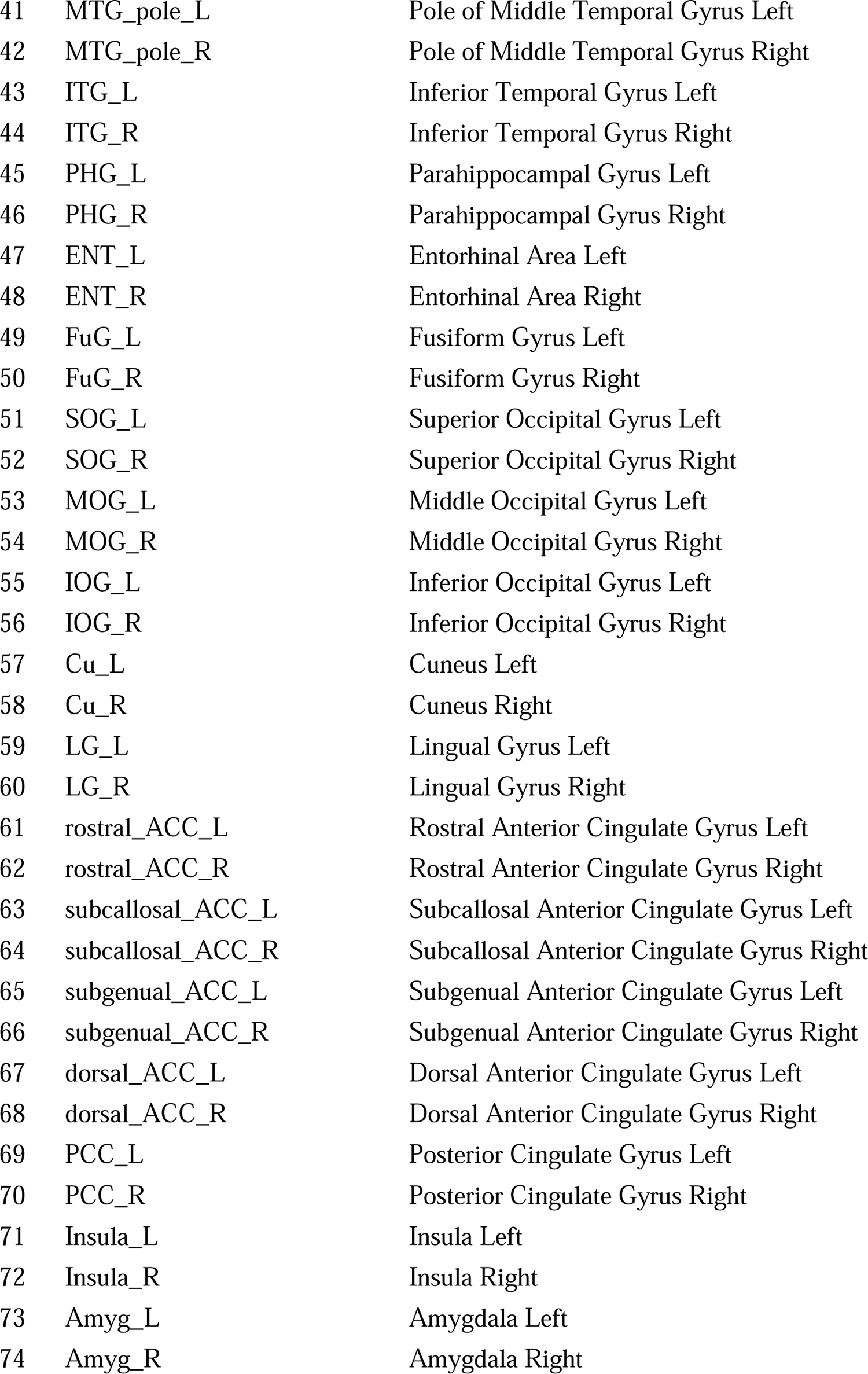
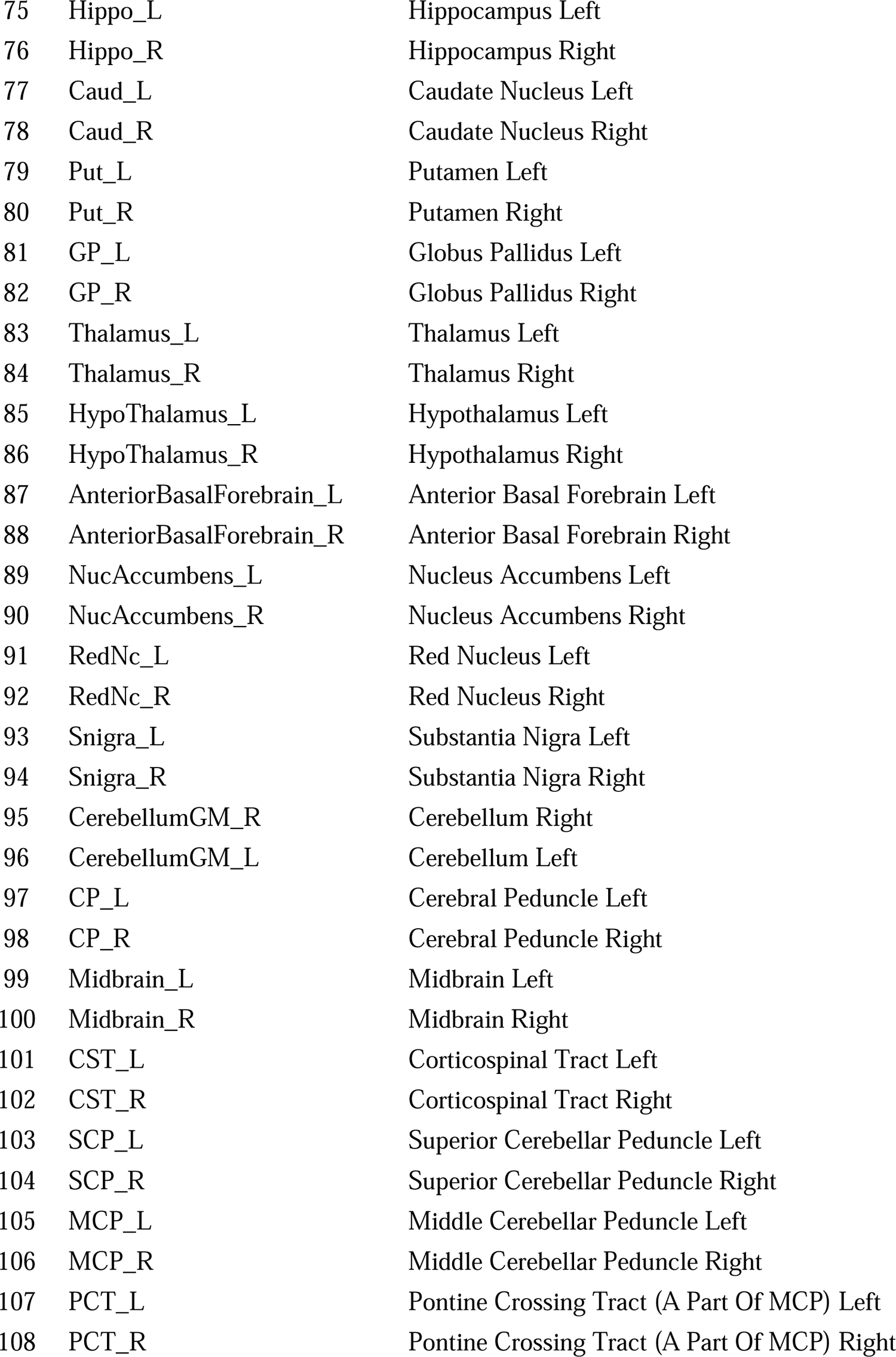
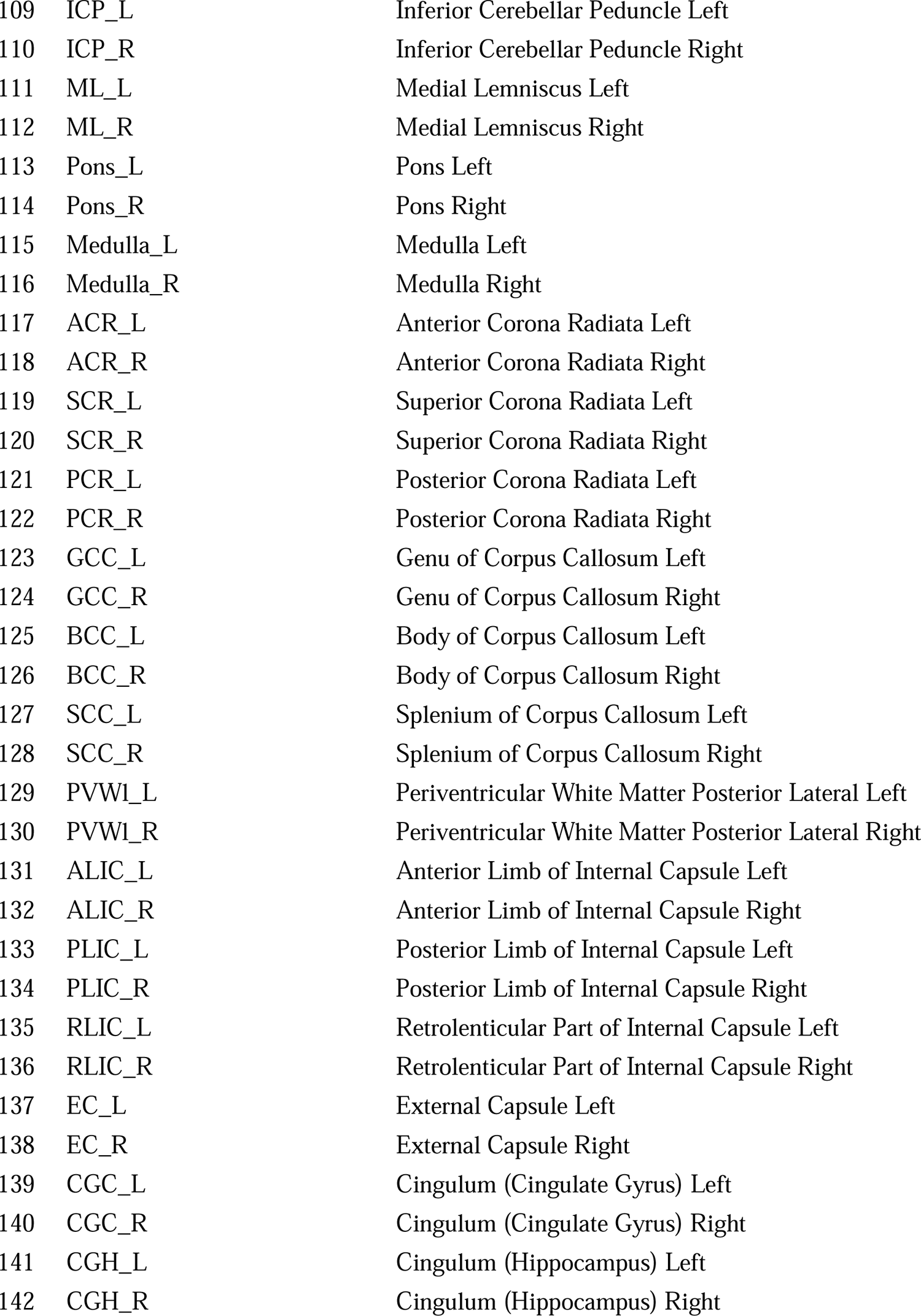
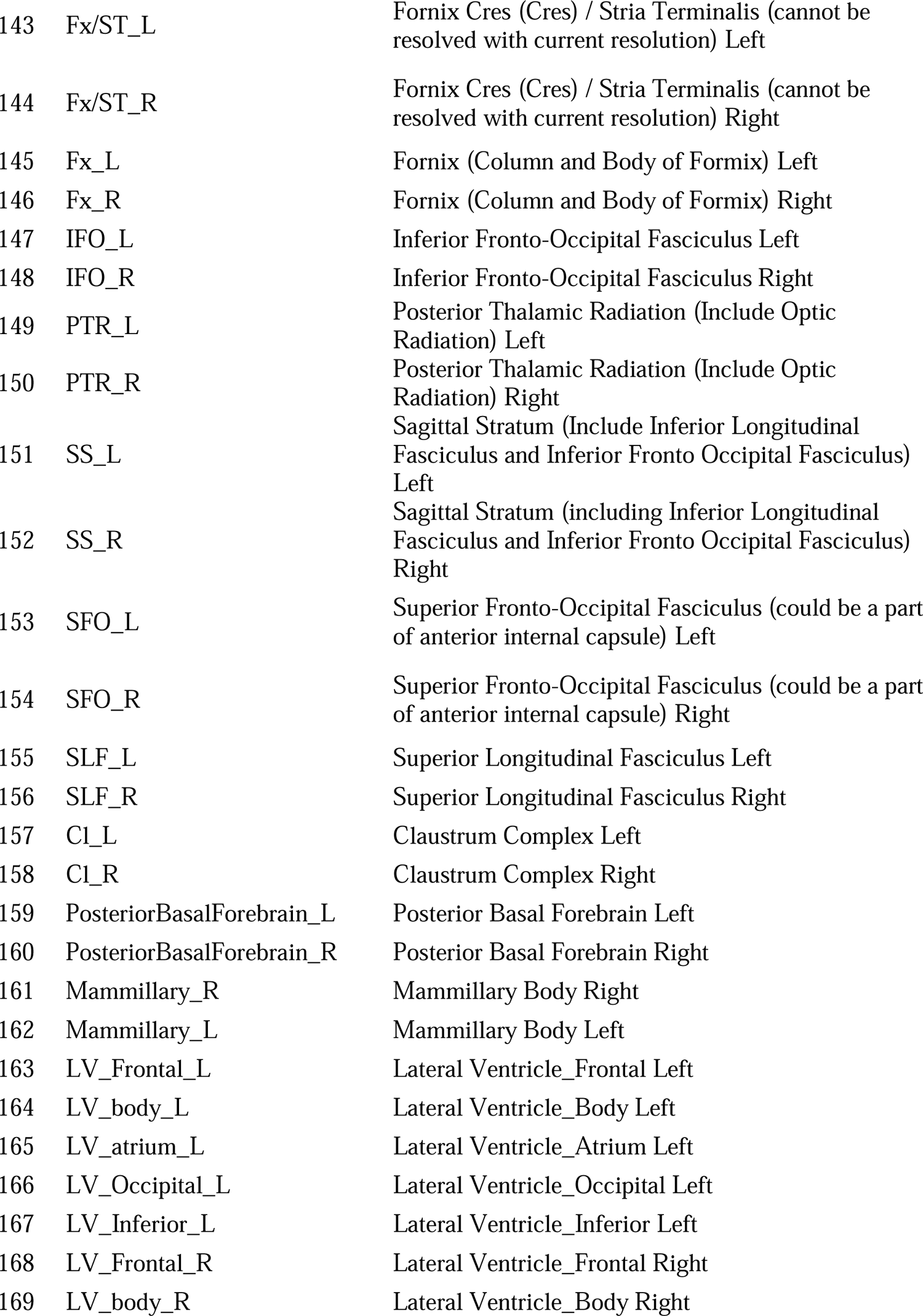
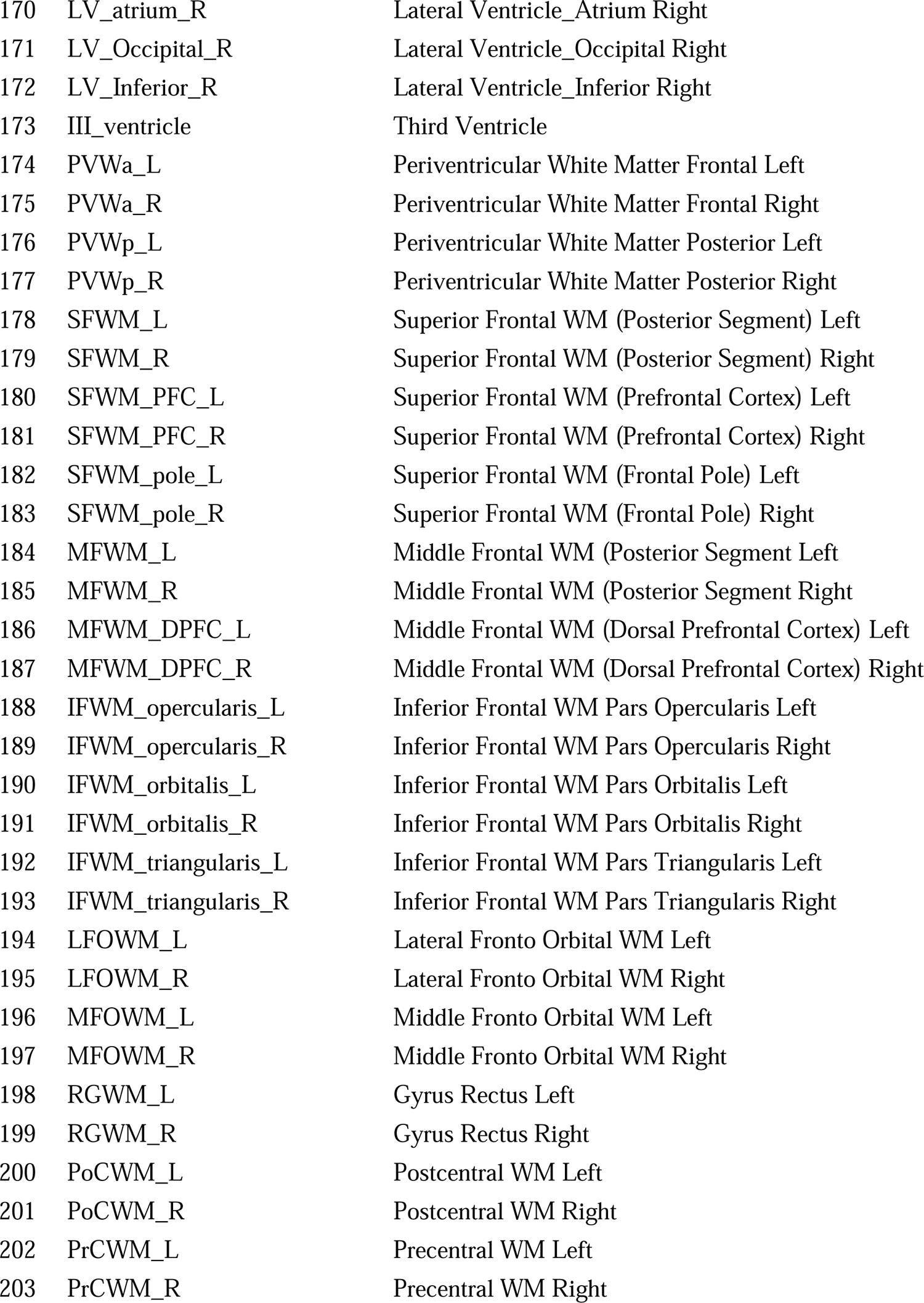
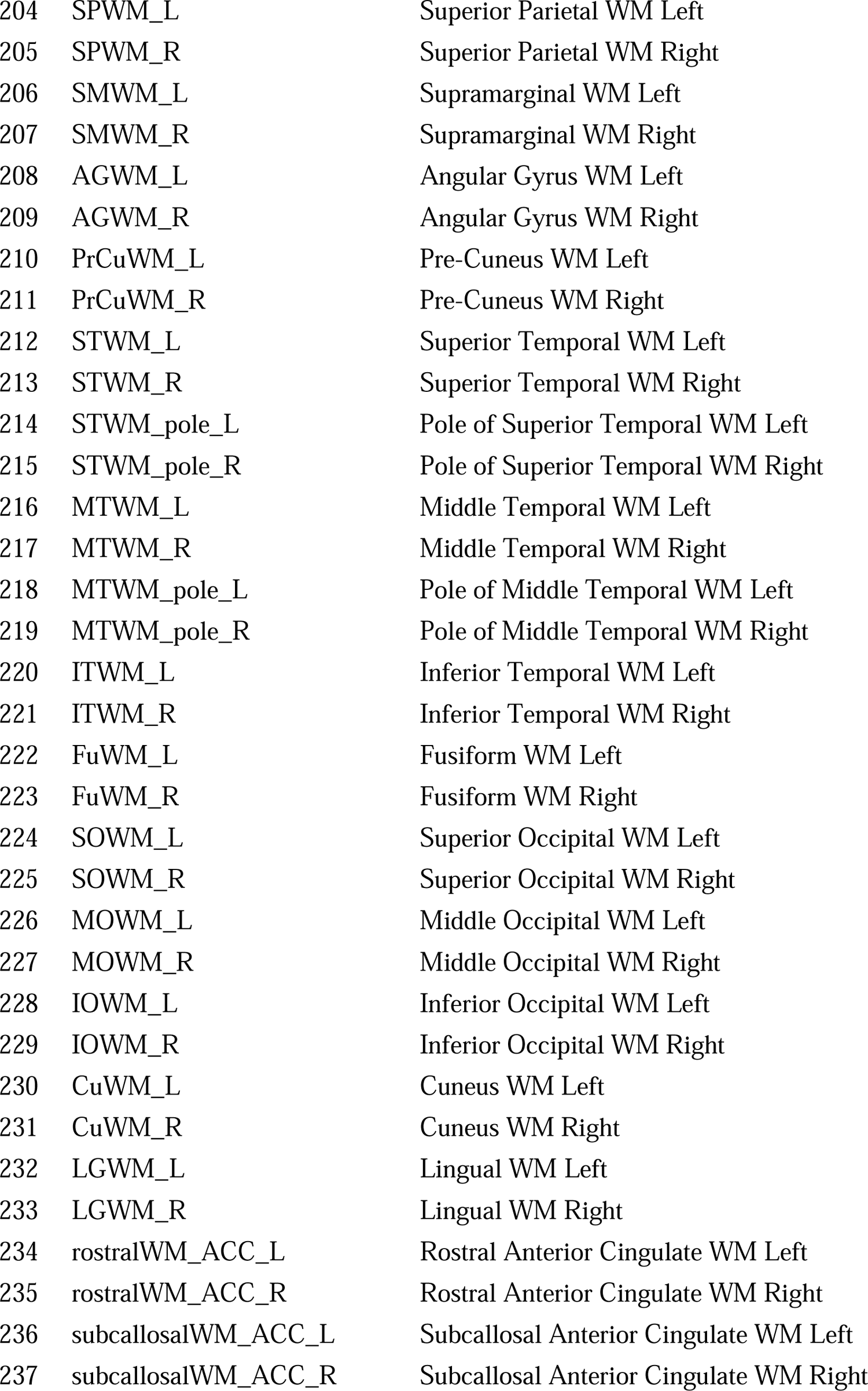
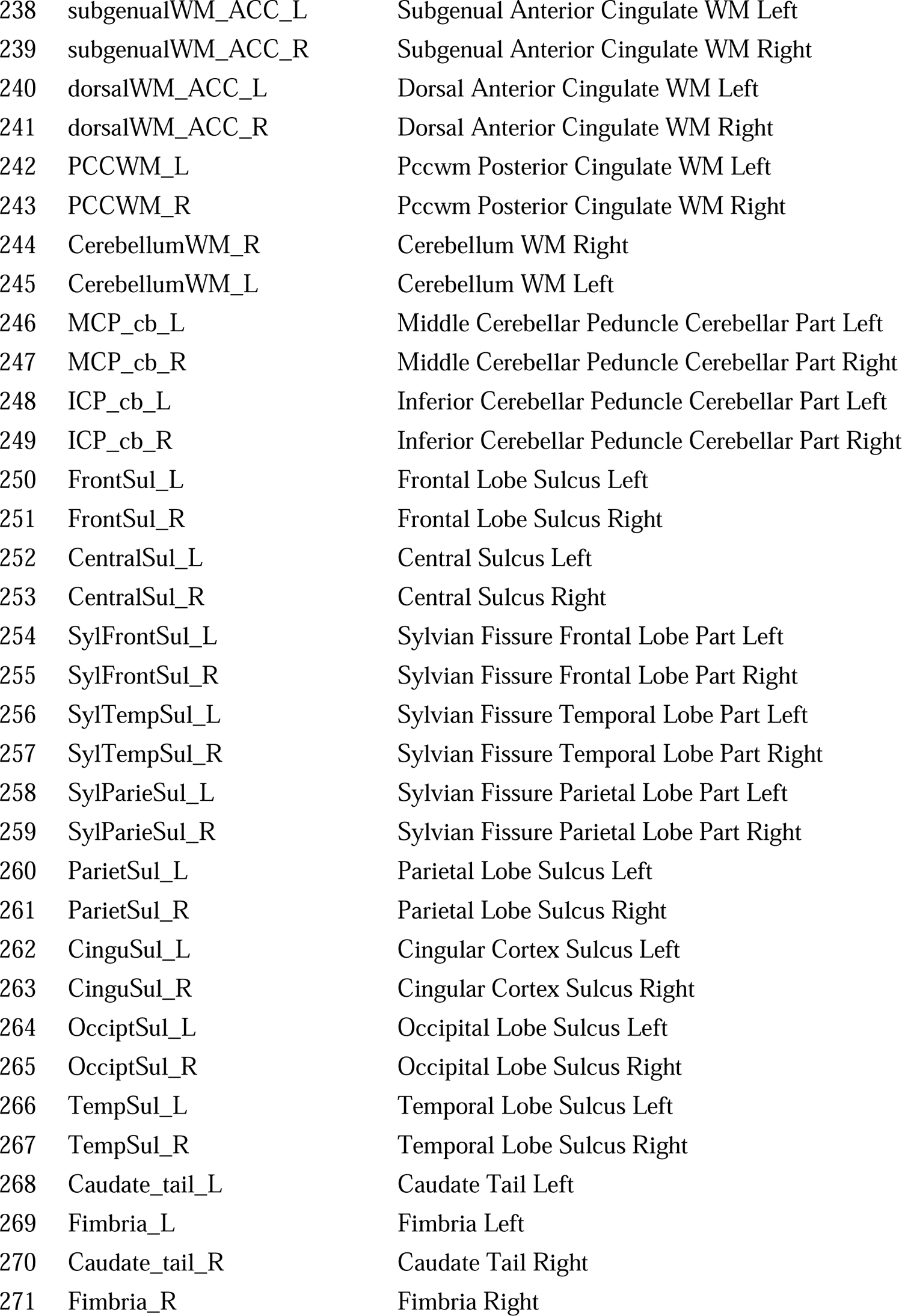
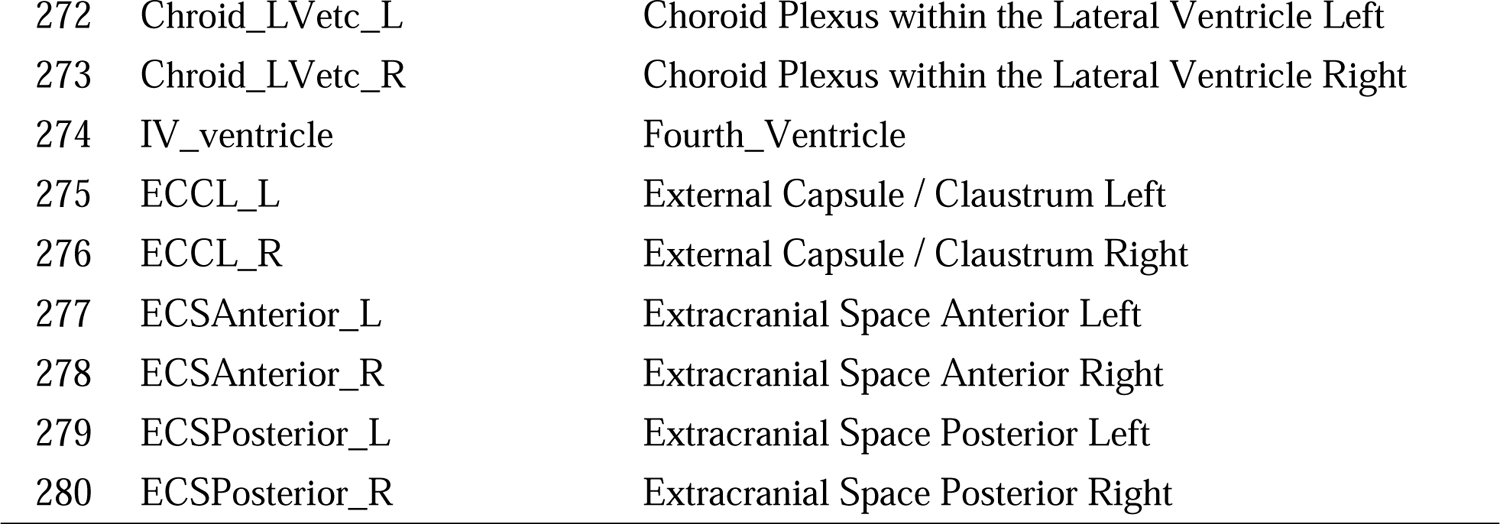
Regions of interest defined in OMAP-T1.

